# Validity, reliability, and readability of single-item and short physical activity questionnaires for use in surveillance: a systematic review

**DOI:** 10.1101/2023.07.19.23292870

**Authors:** Antonina Tcymbal, Sven Messing, Rachel Mait, Roberto Galindo Perez, Taiyeba Akter, Ivo Rakovac, Peter Gelius, Karim Abu-Omar

**Affiliations:** Department of Sport Science and Sport, Friedrich-Alexander-Universität Erlangen-Nürnberg, Gebbertstr. 123b, 91058 Erlangen, Germany; World Health Organization Regional Office for Europe, Marmorvej 51, DK-2100 Copenhagen, Denmark

**Keywords:** physical activity, questionnaire, measurement properties, validity, reliability, readability, surveillance

## Abstract

**Background:** Accurate and fast measurement of physical activity is important for surveillance. Even though many physical activity questionnaires (PAQ) are currently used in research, it is unclear which of them is the most reliable, valid, and easy to use. This systematic review aimed to identify existing brief PAQs, describe and compare their measurement properties, and assess their level of readability.

**Methods:** We performed a systematic review based on the PRISMA statement. Literature searches were conducted in six scientific databases in March 2022. Articles were included if they evaluated validity and/or reliability of brief (i.e., with a maximum of three questions) physical activity or exercise questionnaires intended for healthy adults. Due to the heterogeneity of studies, data were summarized narratively. The level of readability was calculated according to the Flesch-Kincaid formula.

**Results:** In total, 34 articles published in English or Spanish were included, evaluating 31 distinct brief PAQs. The studies indicated moderate to good levels of reliability for the PAQs. However, the majority of results showed weak validity when validated against objective measurements and demonstrated weak to moderate validity when validated against other PAQs. Most of the assessed PAQs met the criterion of being "short," allowing respondents to complete them in less than one minute either by themselves or with an interviewer. However, only 17 questionnaires had a readability level that indicates that the PAQ is easy to understand for the majority of the population.

**Conclusions:** This review identified a variety of brief PAQs, but most of them were evaluated in only a single study. The methods used to assess measurement properties varied widely across studies, limiting the comparability between different PAQs and making it challenging to identify a single tool as the most suitable. Furthermore, PAQs employed different concepts for measuring physical activity, necessitating consideration of measurement properties and assessment goals when selecting a specific tool. None of the evaluated brief PAQs allowed for the measurement of whether a person fulfills the main WHO physical activity recommendations. Future development or adaptation of PAQs should prioritize readability as an important factor to enhance their usability.

## Background

It has been demonstrated that regular physical activity (PA) can help improve physical and mental functions as well as reverse some effects of chronic diseases (1). Regularly engaging in 150 minutes of PA per week (2), alongside following a healthy diet and abstaining from smoking and alcohol consumption, is seen as being key to prevent non-communicable diseases.

However, accurate measurement of PA levels is important to determine the amount of activity needed to improve health and identify links with other health outcomes and behaviors (3). To this day, self-report questionnaires are the most common measures to collect PA data, often as part of surveillance systems such as the Behavioral Risk Factor Surveillance System (4), the WHO STEPwise approach to noncommunicable disease risk factor surveillance (5), and the European Health Interview Survey (6). While such self-report questionnaires have seen widespread use because of their efficiency, they have also been shown to have limitations related to bias and data accuracy (7).

Many different self-report questionnaires have been developed over the years with the Global Physical Activity Questionnaire (GPAQ), the International Physical Activity Questionnaire (IPAQ; also available as a short form, IPAQ-SF), and the European Health Interview Survey Physical Activity Questionnaire (EHIS-PAQ) being the most widely utilized in global PA surveillance (8). All three questionnaires assess PA across different domains, asking respondents to report their PA in a typical week (GPAQ, EHIS-PAQ) or during the last 7 days (IPAQ-SF). All three are comparatively complex and range from seven items (IPAQ-SF) to 16 (GPAQ). Nevertheless, the measurement properties of these questionnaires are modest. For GPAQ (9), IPAQ-SF (10, 11), and the EHIS-PAQ (12), different studies have demonstrated reasonable reliability but comparatively low validity. In a recent review (8) of IPAQ-SF, GPAQ, and EHIS-PAQ, the questionnaires showed low-to-moderate validity against objective measures of PA such as accelerometers, and moderate-to-high validity against subjectively measured PA such as other questionnaires.

It is well known that questionnaires with many items can increase the response burden on respondents (13, 14), which has resulted in the development of several short self-report instruments. In contrast to the detailed questionnaires mentioned above, the purpose of short PA questionnaires is to simplify and speed up the procedure for assessing PA levels.

In addition, language-related difficulties are currently an important topic in public health. Patient education materials can increase patient compliance, but only if they are written in a language that is easy for the patient to understand (15). Regarding PA questionnaires, Altschuler et al. (16) found significant gaps between respondents’ interpretations of some PA questions and researchers’ original assumptions about what those questions were intended to measure. One of the characteristics of language difficulty is the level of readability, which indicates how easily readers can understand the text. Research of texts used in healthcare consistently shows that materials intended for patients often require a high level of education and are too complicated for the average person (15, 17). In relation to physical activity questionnaires (PAQs), readability can influence the amount of time a person needs to understand the question and, if the text is too complicated, may potentially decrease the response rate and correctness of the answer.

A number of existing reviews have investigated the measurement properties of PA questionnaires. Van Poppel et al. (18) reviewed the validity and reliability methodology of 85 versions of PAQs with no clear consensus regarding the best questionnaire for PA measurement. Helmerhost et al. (19) studied reliability and objective criterion-related validity of 34 newly-developed and 96 existing PAQs.

However, to our knowledge, a dedicated review of short and brief PA questionnaires measurement properties has not yet been performed. Therefore, the aim of this systematic review is to identify existing brief PAQs and to comparatively describe their measurement properties and level of readability.

## Methods

This review follows the guidelines of the PRISMA (Preferred Reporting Items for Systematic reviews and Meta-Analyses) statement (20) and the Consensus-based Standards for the Selection of Health Measurement Instruments (COSMIN) guideline for systematic reviews of patient-reported outcome measures (21).

### Information sources and search strategy

A systematic search was performed in six databases (PubMed, Web of Science, Scopus, CINAHL, SportDiscus, PsycInfo) in March 2022. Additionally, reference lists from previous reviews of PA questionnaires and other relevant publications were screened to identify additional studies. A comprehensive search strategy was developed with a combination of keywords in the categories of measured construct (physical activity) and type of instruments (brief/short questionnaire).

The resulting search string was as follows:

("physical activit*" OR "physical inactivit*")

AND

(questionnaire OR measure OR evaluat* OR assess* OR surveillance OR monitor* OR screening)

AND

("single item" OR single-item OR "single question" OR "one item" OR one-item OR "one question" OR "brief questionnaire " OR "short questionnaire " OR "brief assessment" OR "short assessment" OR "two-item*" OR "two-item*" OR "two questions" OR "brief physical activity assessment" OR "Single response" OR "Single-response")

No restrictions were made regarding language or publication date.

### Eligibility criteria

Articles were included in the review if they fulfilled the following eligibility criteria:

1. The article described a self-report PA or exercise questionnaire intended for healthy adults.
2. The evaluated questionnaire was brief and included a maximum of three questions.
3. The article investigated one or more measurement properties of the questionnaire.
4. The article was published in a peer-reviewed journal.

Articles were excluded based on the following exclusion criteria:

1. The article focused on a questionnaire that measured only physical inactivity, screen time, or sedentary behavior.
2. The evaluated questionnaire was intended solely for children and adolescents or people with specific conditions.
3. The article did not investigate the measurement properties of the questionnaire.

### Study selection

Two reviewers independently screened and selected the relevant articles. First, all articles were screened based on titles and abstracts. If the title and/or abstract indicated that the study fulfilled the inclusion criteria, both reviewers screened the full text for eligibility. When necessary, supplementary files were also reviewed for additional information. Disagreements between the reviewers were discussed within the research team until a consensus was reached.

Records were managed using the Covidence systematic review software (Veritas Health Innovation, Melbourne, Australia; www.covidence.org) and EndNote X9 (Clarivate Analytics, Philadelphia, PA, USA).

### Data extraction

Data of included studies were extracted and summarized by one reviewer and verified by a second reviewer to reduce bias and error. Discrepancies were discussed between reviewers to achieve consensus. Extracted information included: publication details (first author, year of publication, country), sample characteristics (number of participants, age category, special health conditions), the measurement tool(s) explored, who assessed PA levels of participants, assessed measurement properties, other measurement tool(s) used as a comparison, reliability test-retest interval, and the results of the study.

### Risk of bias assessment

The methodological quality of the individual studies included in this review was assessed with 18 questions based on the Appraisal tool for Cross-Sectional Studies (AXIS) (22). In order to specifically assess the quality of the tools’ validity and reliability measurement properties, some questions were modified based on the COSMIN risk of bias checklist (21) and on suggestions from previous systematic reviews examining PA assessment measures (23). This quality assessment evaluated articles based on study design, sample size, participant selection process, appropriate blinding, examiner experience, method of measurement, adequate data reporting, internal consistency, and six other categories (see Additional file 1). Risk of bias assessment was conducted independently by two reviewers, with any discrepancies resolved through discussion in the research team. Studies were scored 1 if they satisfied the quality element, and 0 if they did not. The summary score (range: 0–18) indicates the risk of bias, with a higher score indicating higher quality and therefore a lower risk of bias.

### Data synthesis and analysis of measurement properties

The primary objective of this systematic review was to identify existing short questionnaires suitable for assessing PA levels in surveillance and primary care settings, and to compare their measurement properties. To accomplish this goal, relevant information from the included studies was summarized separately for each questionnaire. In order to evaluate the reliability and validity of the identified questionnaires, a range of tests were employed in the included studies. Some studies reported results for a total questionnaire summary score, while others assessed reliability and validity for specific aspects, intensities, or domains of the questionnaire. Additionally, certain studies examined these measurement properties within subgroups of the test population. Due to the heterogeneity in the methods used and the lack of standardized reporting across studies, a quantitative meta-analysis was not feasible. Consequently, the information from the included studies was summarized narratively, highlighting the key findings for each questionnaire.

This narrative synthesis allows for a comprehensive overview of the reliability and validity findings, highlighting strengths and limitations of each questionnaire.

### Analysis of length and readability of questionnaires

To determine how quickly questionnaires could be answered and how easy it is to understand the questions, identified PAQs were analyzed for word count and readability level. The expected time the tool would take for self-administration (silent reading speed) and interviewer administration (spoken word speed) was calculated based on the respective questionnaire’s word count and English reading speeds established by Brysbaert (24). The level of readability was calculated according to the Flesch-Kincaid formula, which was chosen because it is the most commonly used tool to calculate the readability level of written health information (25). The formula has two forms: the Flesch Reading-Ease-Score, and the Flesch–Kincaid Grade Level. The Flesch Reading-Ease-Score test produces a score from 0 to 100, and higher scores indicate material that is easier to read; lower scores mark passages that are more difficult to read. The Flesch–Kincaid Grade Level formula matches the text to the grade level achievement (number of years of education) required to understand the text. We applied both formulas for each questionnaire using an online calculator (26).

## Results

### Study selection process

The search across six databases resulted in 2,422 publications. After removing 1,255 duplicates, 1,167 articles were screened based on title and abstract. 65 studies were found eligible for full text assessment. One recently published article (27) and one article identified while screening the reference lists of relevant publications (28) satisfied all eligibility criteria and were therefore included in the analysis.

Subsequently, 33 full text articles were excluded due to the length of the questionnaire (n=14), the publication type (n=14), a lack of self-reporting on PA and exercise (n=2), and a study design that did not consider validity and reliability (n=3). In total, 34 studies were included for data extraction and methodological quality assessment. A summary of the search results is presented in Figure 1.

**Figure 1.**
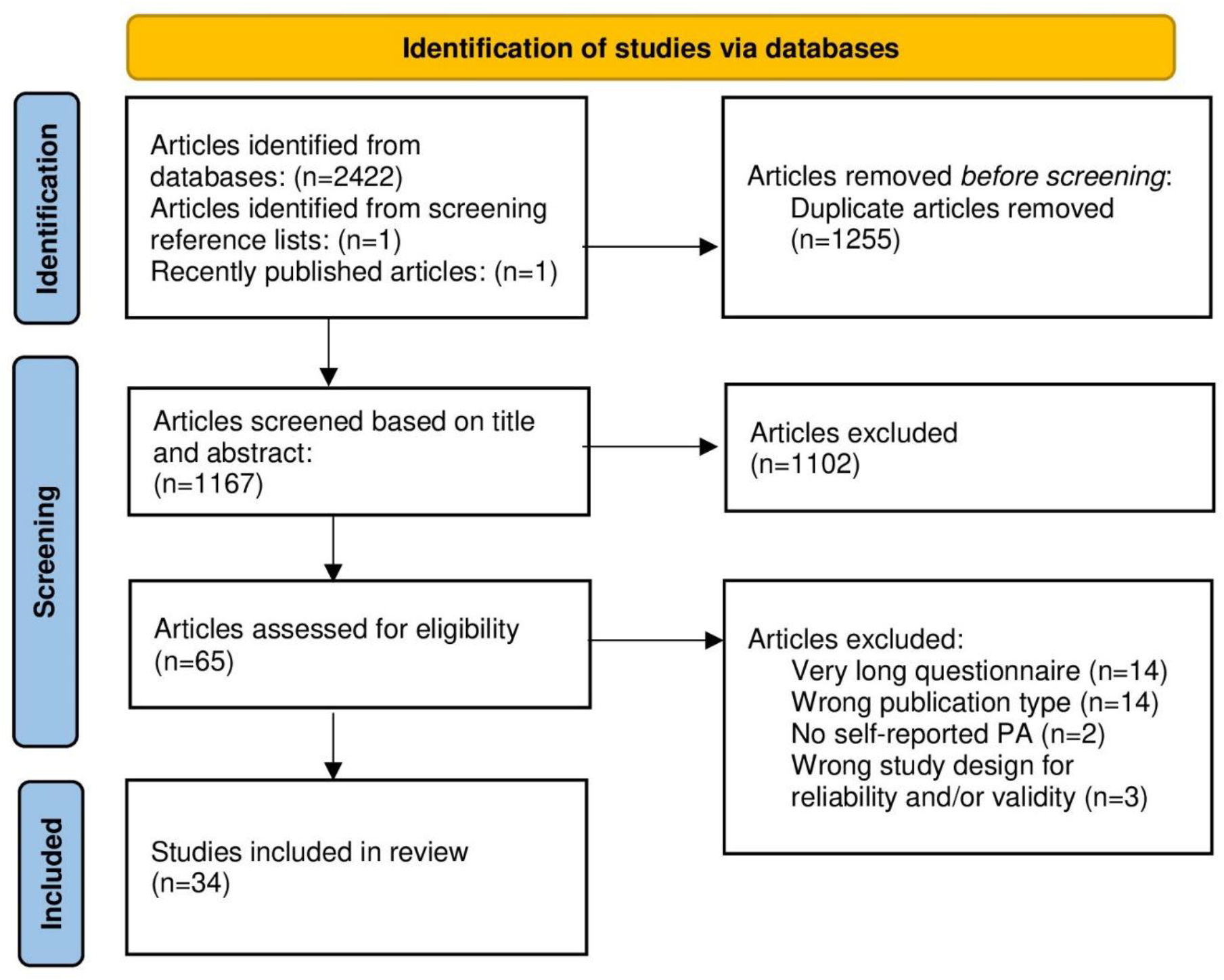
PRISMA flowchart of study eligibility (PA=physical activity)

### Characteristics of included studies

A summary of the characteristics of the 34 included studies is presented in Table 1. The included studies were conducted in Western European countries (n=14), the USA (n=10), Australia (n=5), Canada (n= 2), New Zealand (n=2), and Japan (n=1).

**Table 1.**
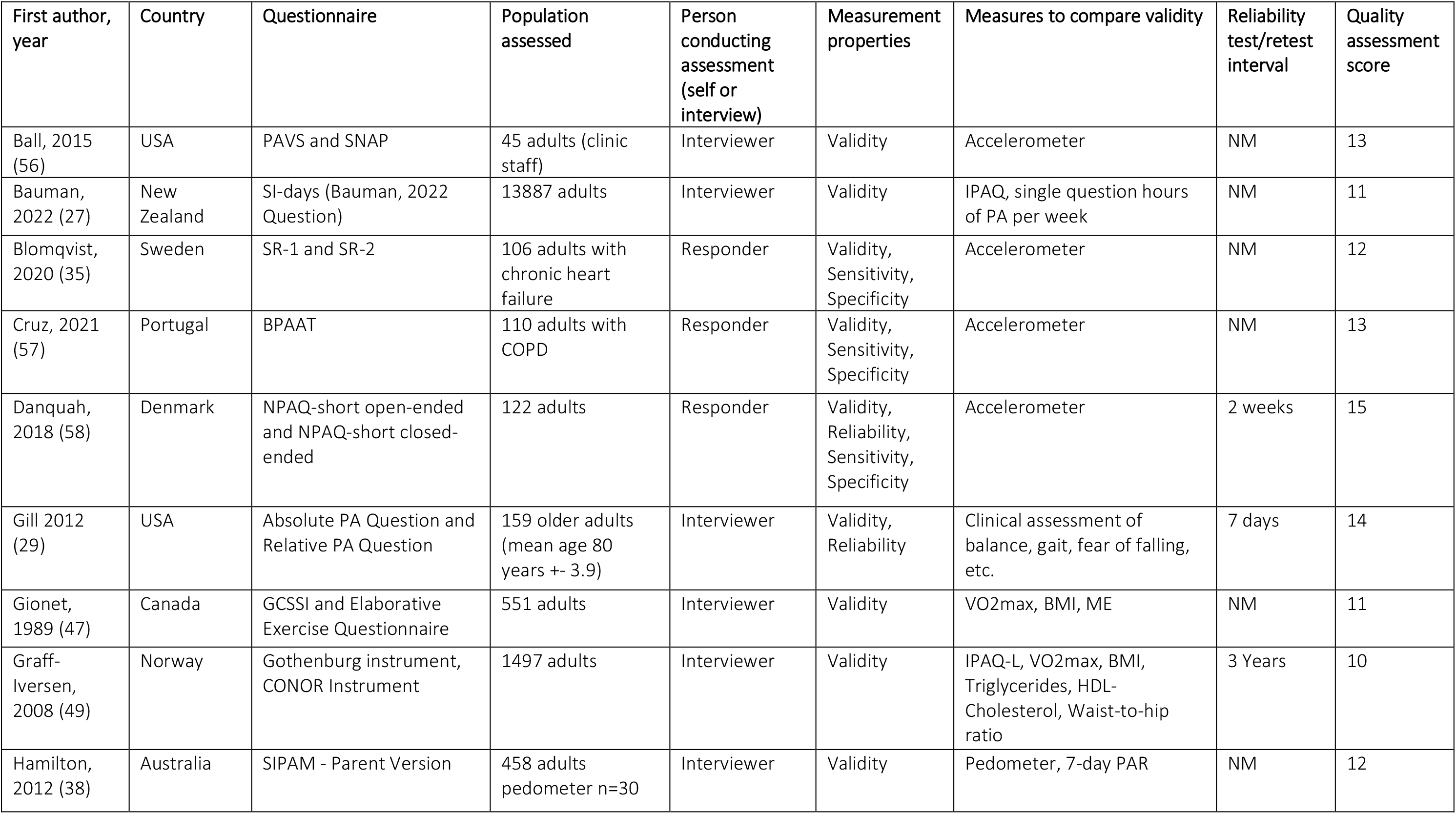
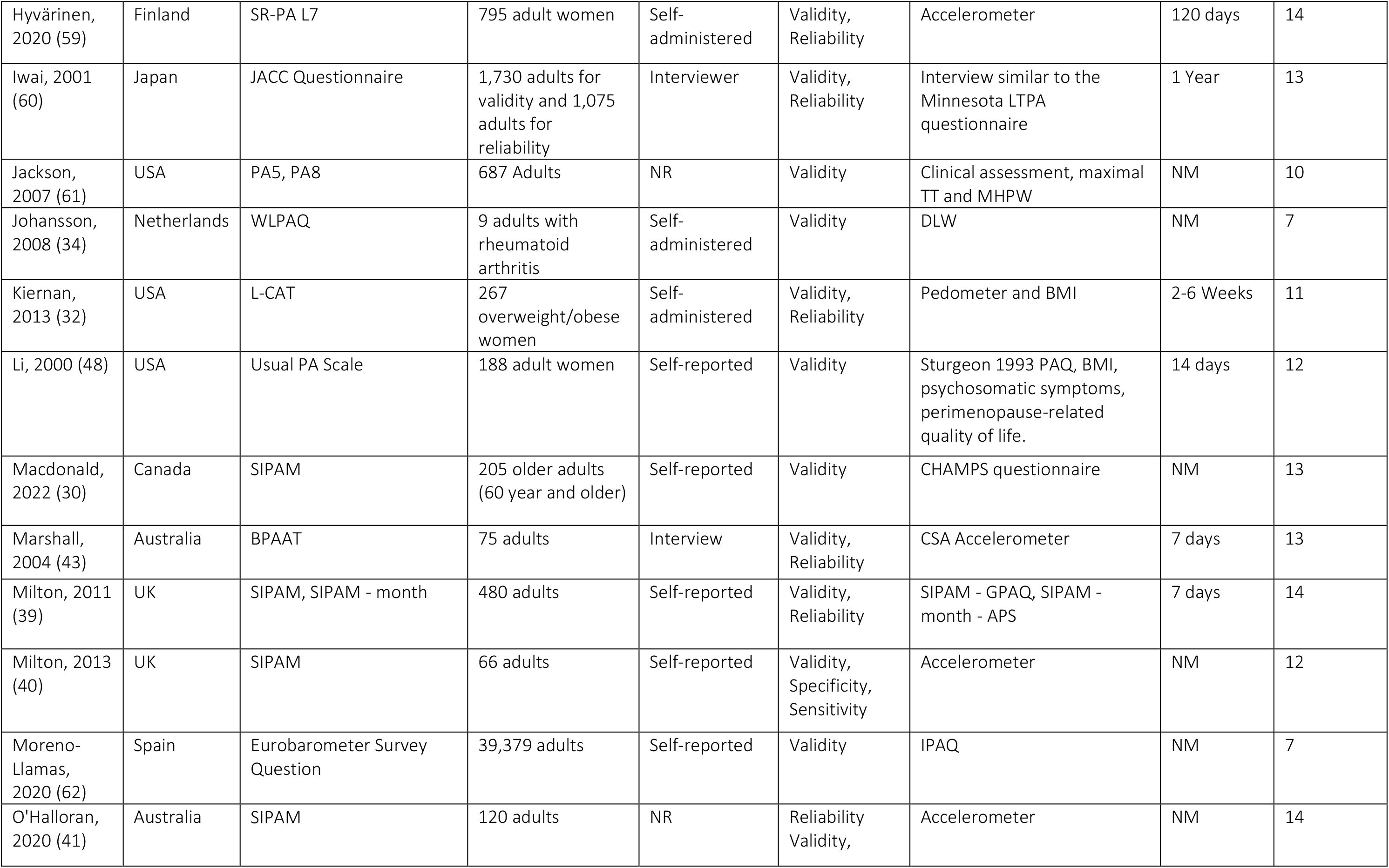
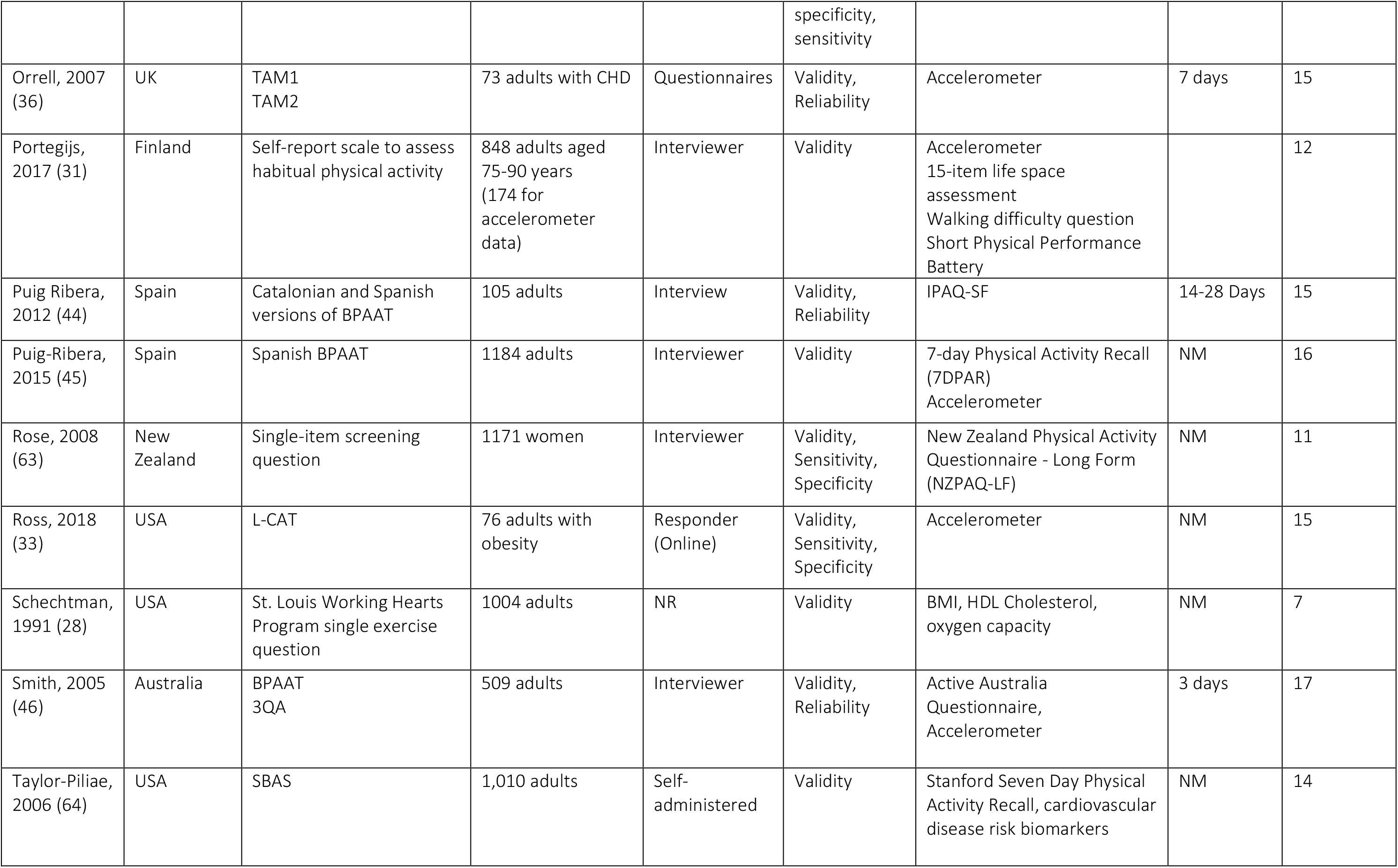
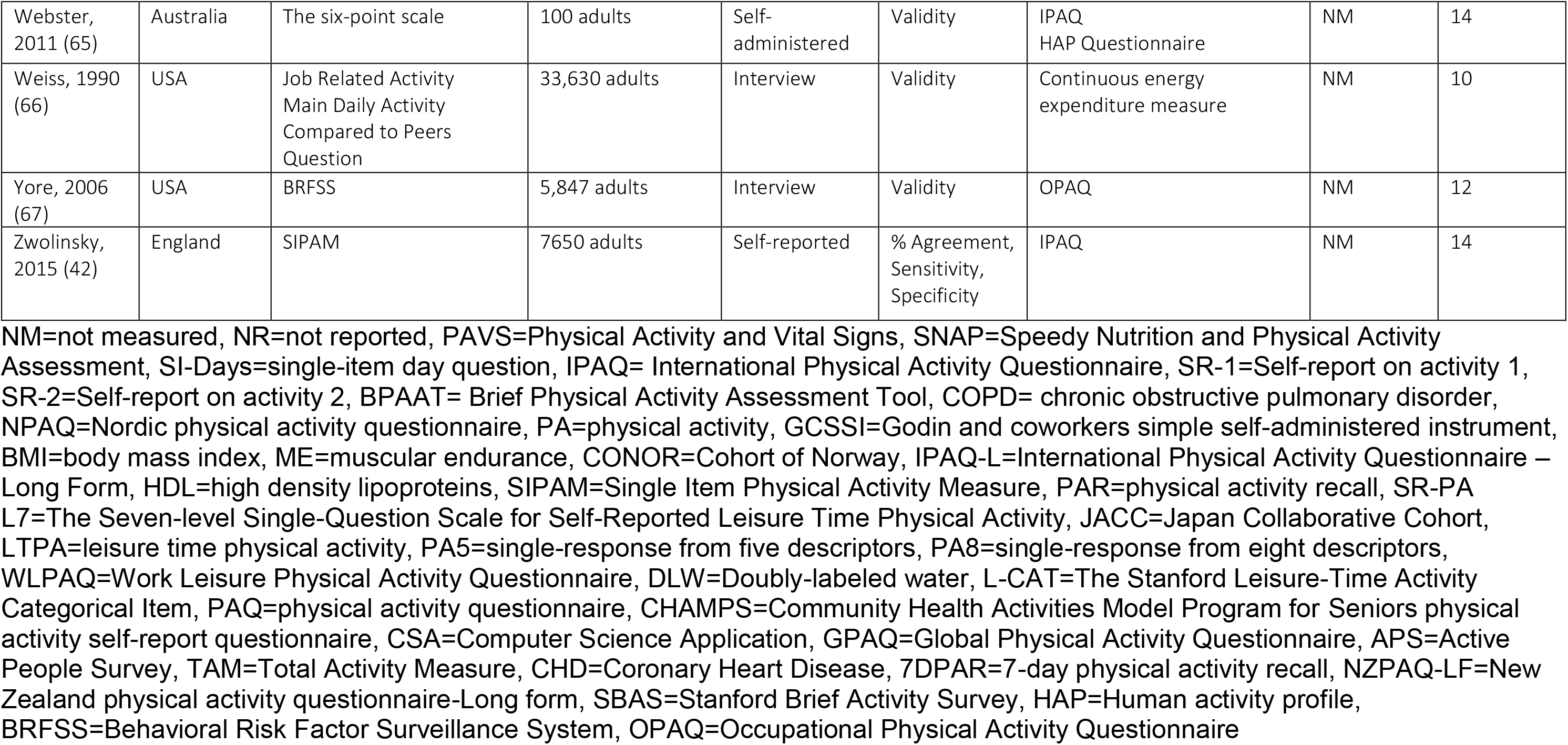
Studies overview.

A total of 114,143 adults were assessed using the 31 unique brief PAQs (with sample sizes ranging from 9 to 39,379). In 25 studies, the sample consisted only of healthy adults, and nine studies also included specific populations such as older adults (29) (30) (31), overweight/obese adults (32, 33), as well as patients with rheumatoid arthritis (34), coronary heart disease (35) (36), and chronic obstructive pulmonary disease (37).

Thirty-one studies documented who completed the PAQ. In 16 studies, respondents filled out the questionnaire by themselves, and in 15 studies the PA level was collected from an interviewer reading a questionnaire.

Questionnaires asked for the amount of PA (n=6), the number of days per week with a sufficient amount of PA (n=4), general exercise participation (n= 3), self-reported activity compared with peers (n=2), or for respondents to choose a categorical descriptor of PA levels ranging from “inactive” to “very active” (n=16).

Seven of the included studies investigated the measurement properties of the Single Item Physical Activity Measure (SIPAM) (27, 30, 38, 39, 40, 41, 42), five were related to the Brief Physical Activity Assessment Tool (BPAAT) (37, 43, 44, 45, 46), and two to the Stanford Leisure-Time Activity Categorical Item (L-Cat) (32, 33). The other 28 PAQs were assessed in a single study.

### Risk of bias assessment

The average quality score of included studies was 11.75, with a range between 7 and 17 (out of 18). Thirteen studies received 14 or more points, which can be considered a high methodological quality. In most of the studies, the aims and objectives, the process of the measurement properties investigation, the statistical analysis and the results were sufficiently described, and the study design was appropriate. However, the included papers reported poorly on whether the examiners had enough experience with the tool and whether they were blinded to participant characteristics, previous findings, or other observed findings. Only four studies justified their choice of sample size. Additionally, many studies failed to consistently report reasons for dropout and characteristics of non-responders. Quality scores of the individual studies are reported in Table 1.

### Questionnaires’ measurement properties

A summary of the reliability, validity, and diagnostic test accuracy data found in the identified studies is presented in Table 2. Studies commonly use a number of different statistical analyses to define absolute (agreement between the two measurement tools) or relative (the degree to which the two measurement tools rank individuals in the same order) validity and reliability. These types of statistical analysis include correlations (Pearson’s; Spearman’s; interclass), regression, kappa statistics, and area under the receiver operating curve (AUC-ROC).

**Table 2.**
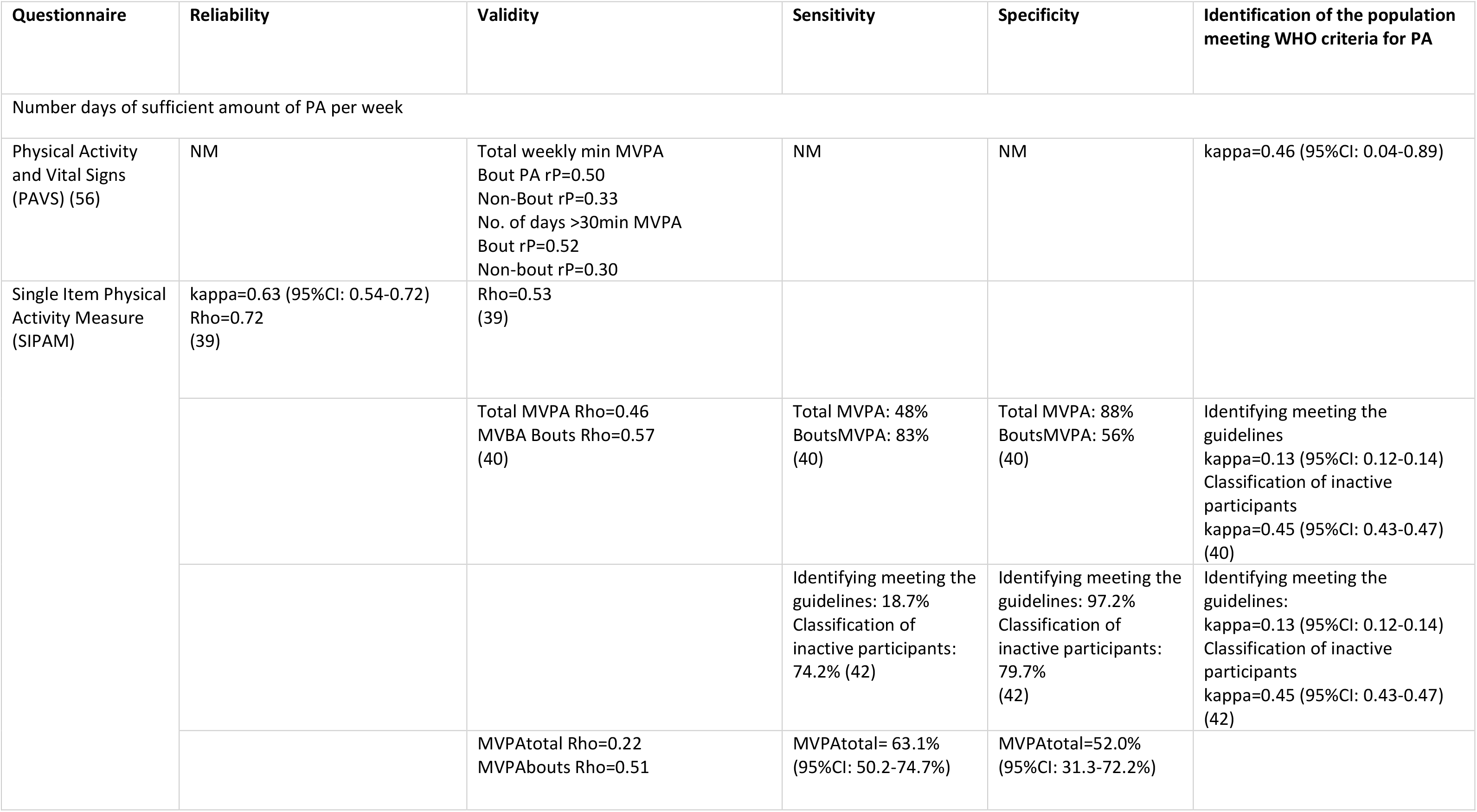
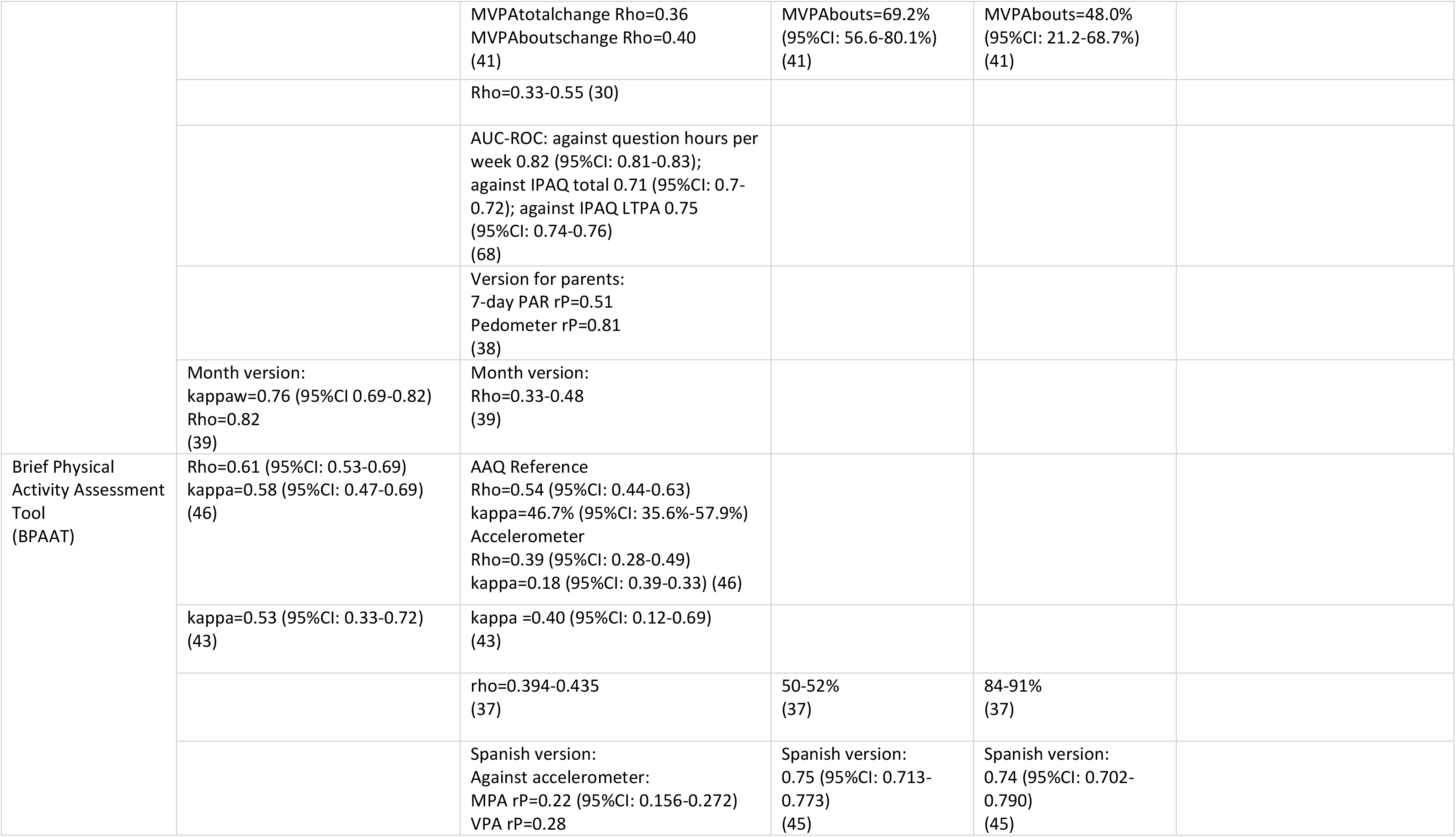
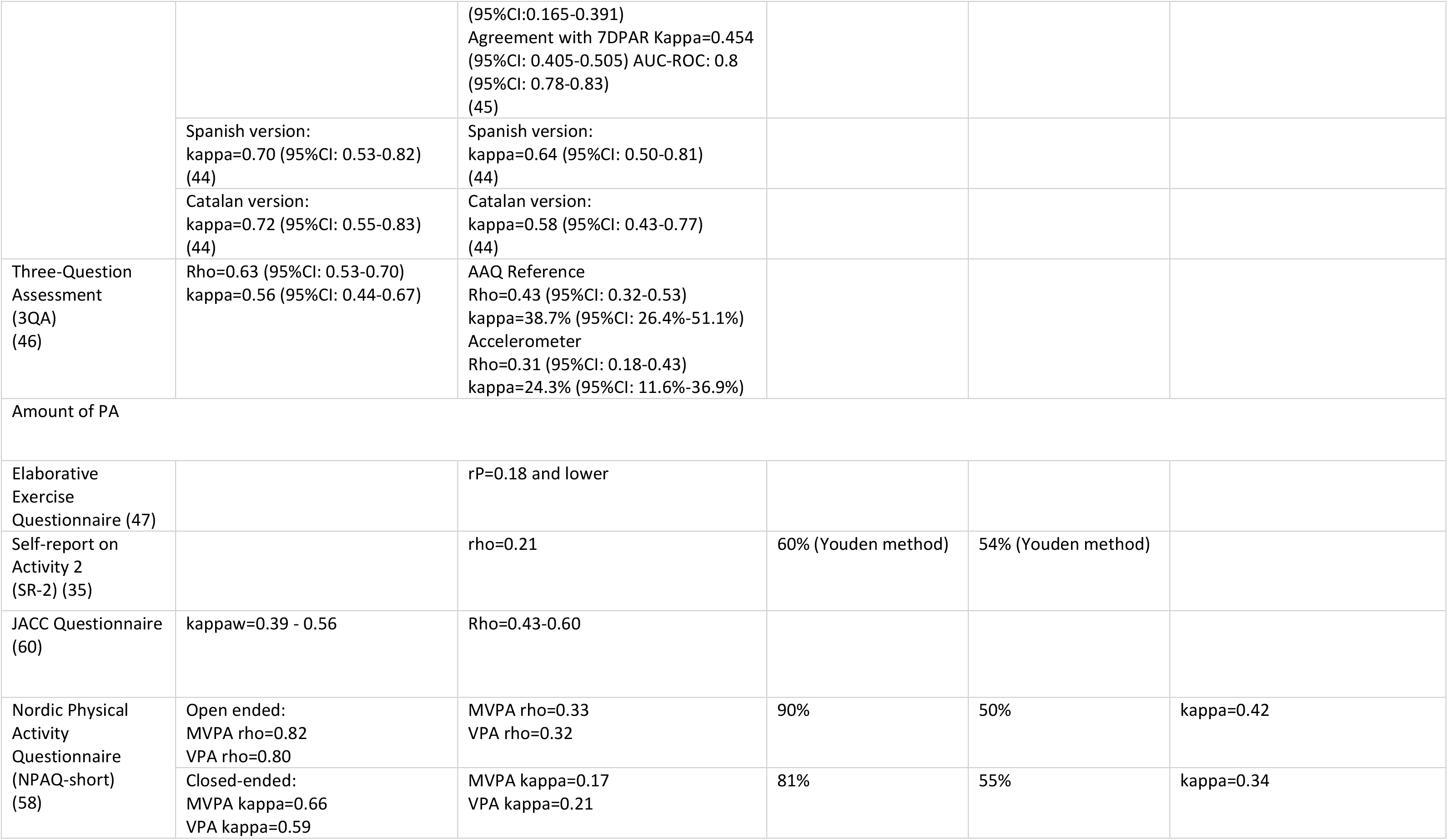
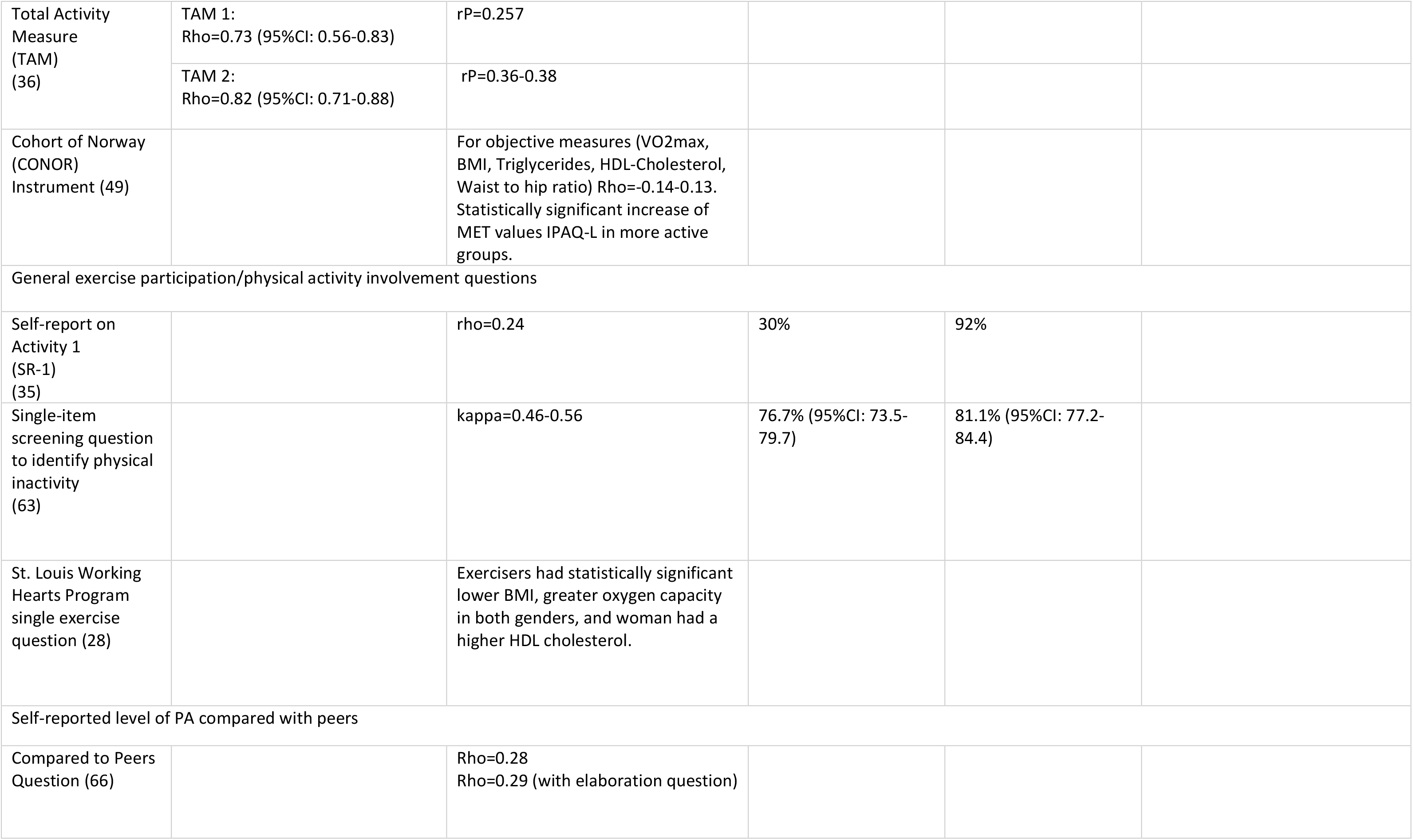
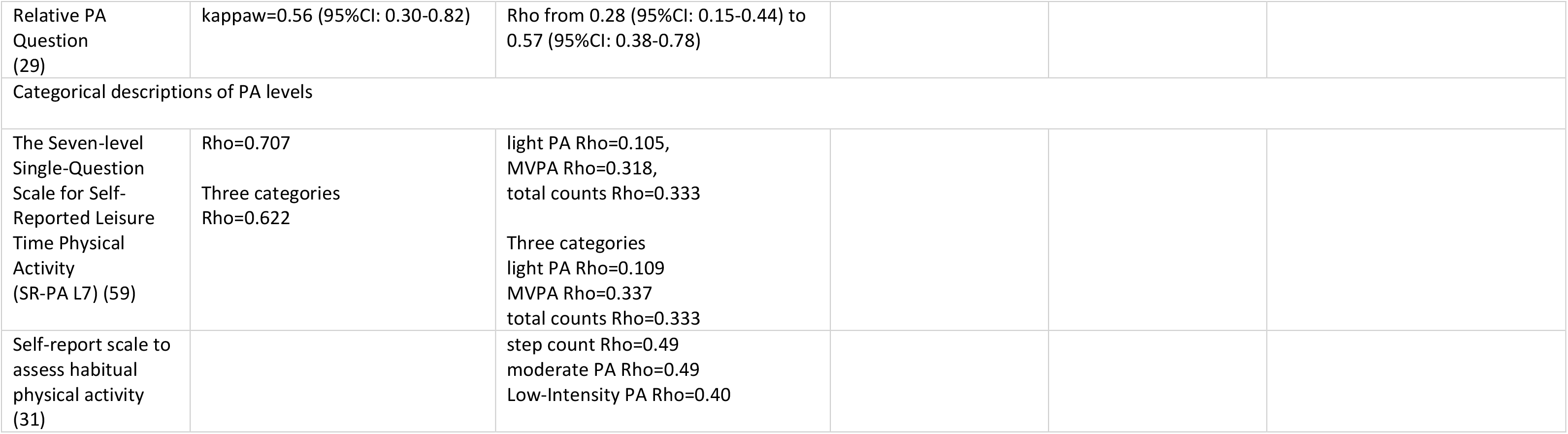
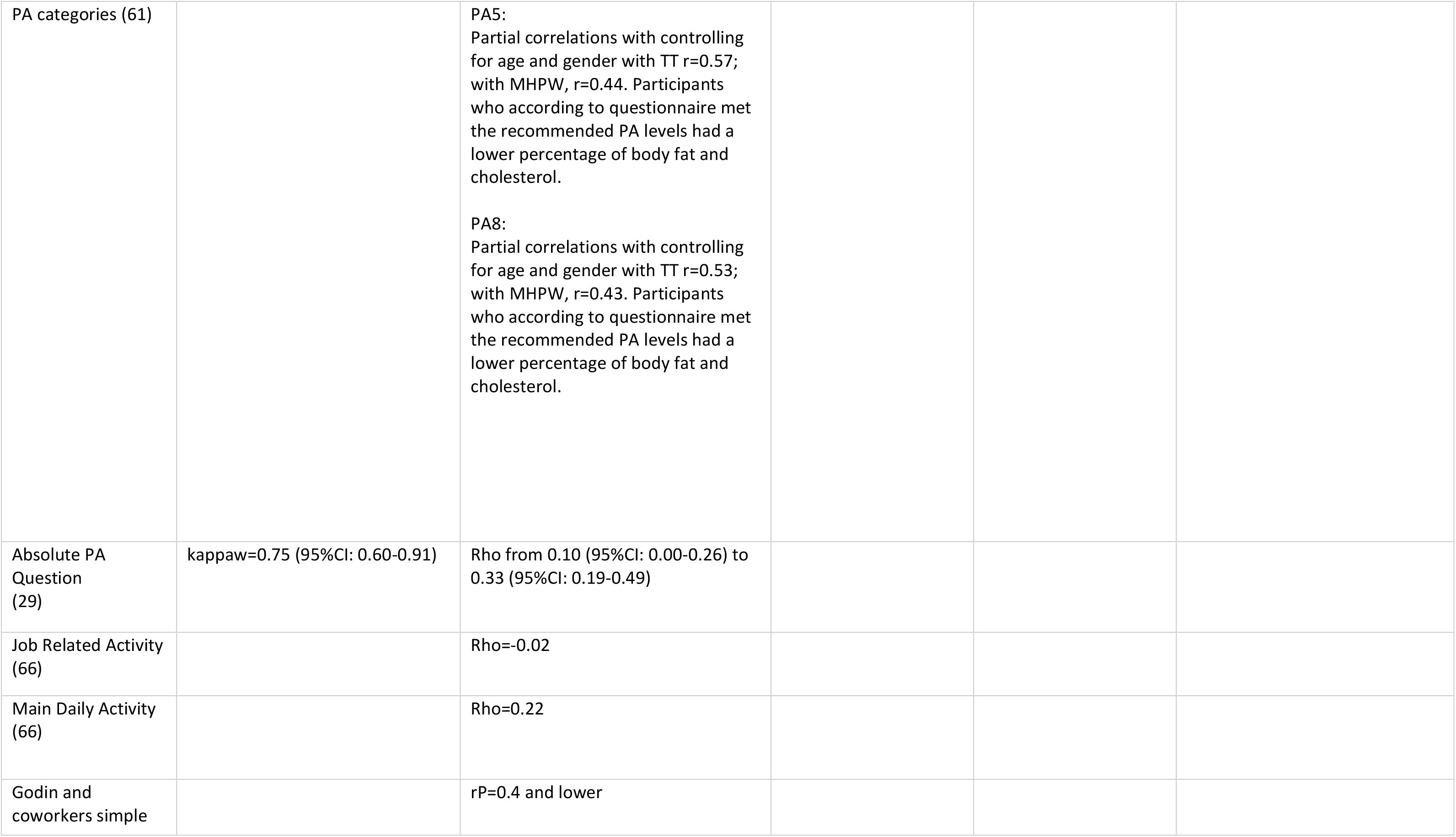
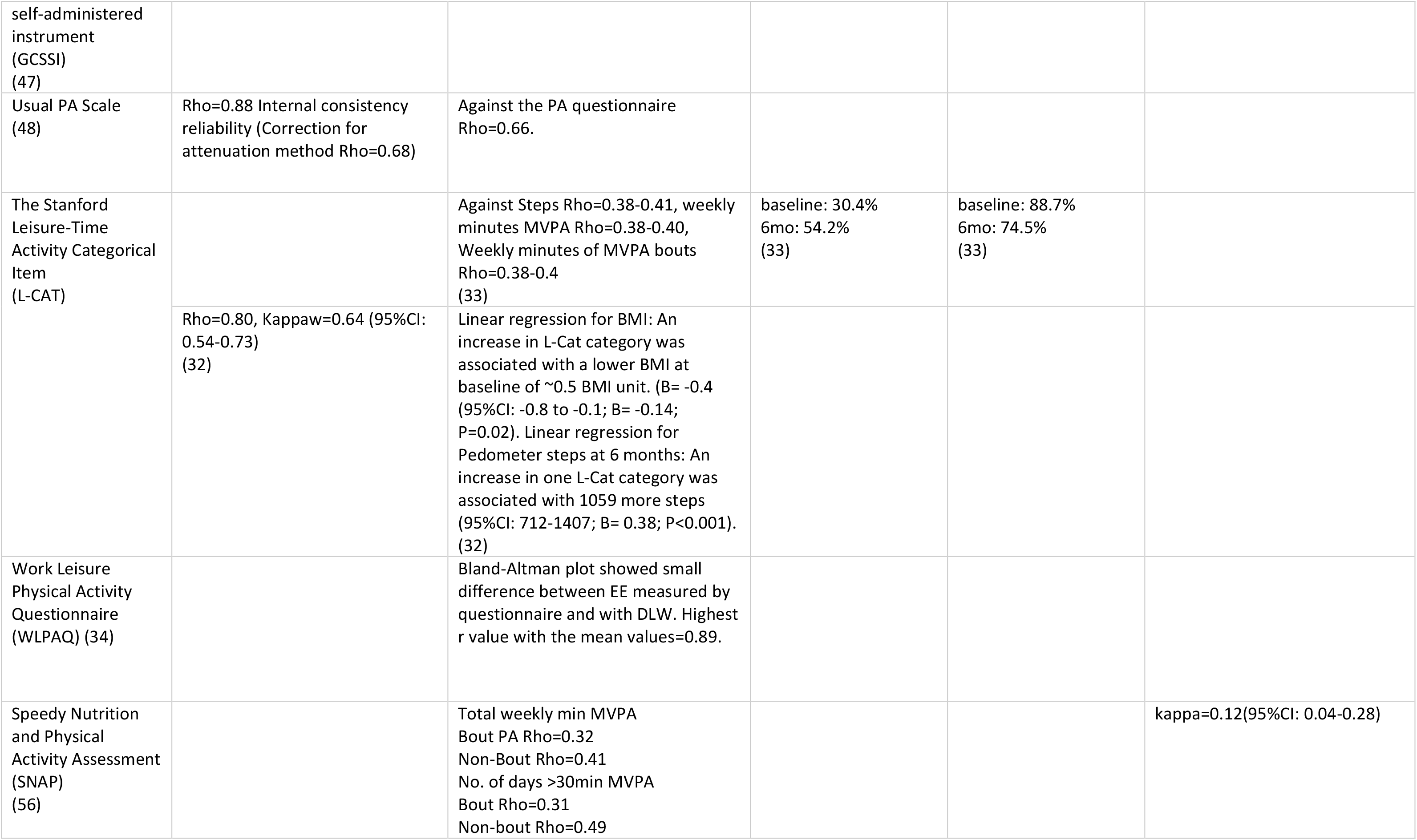
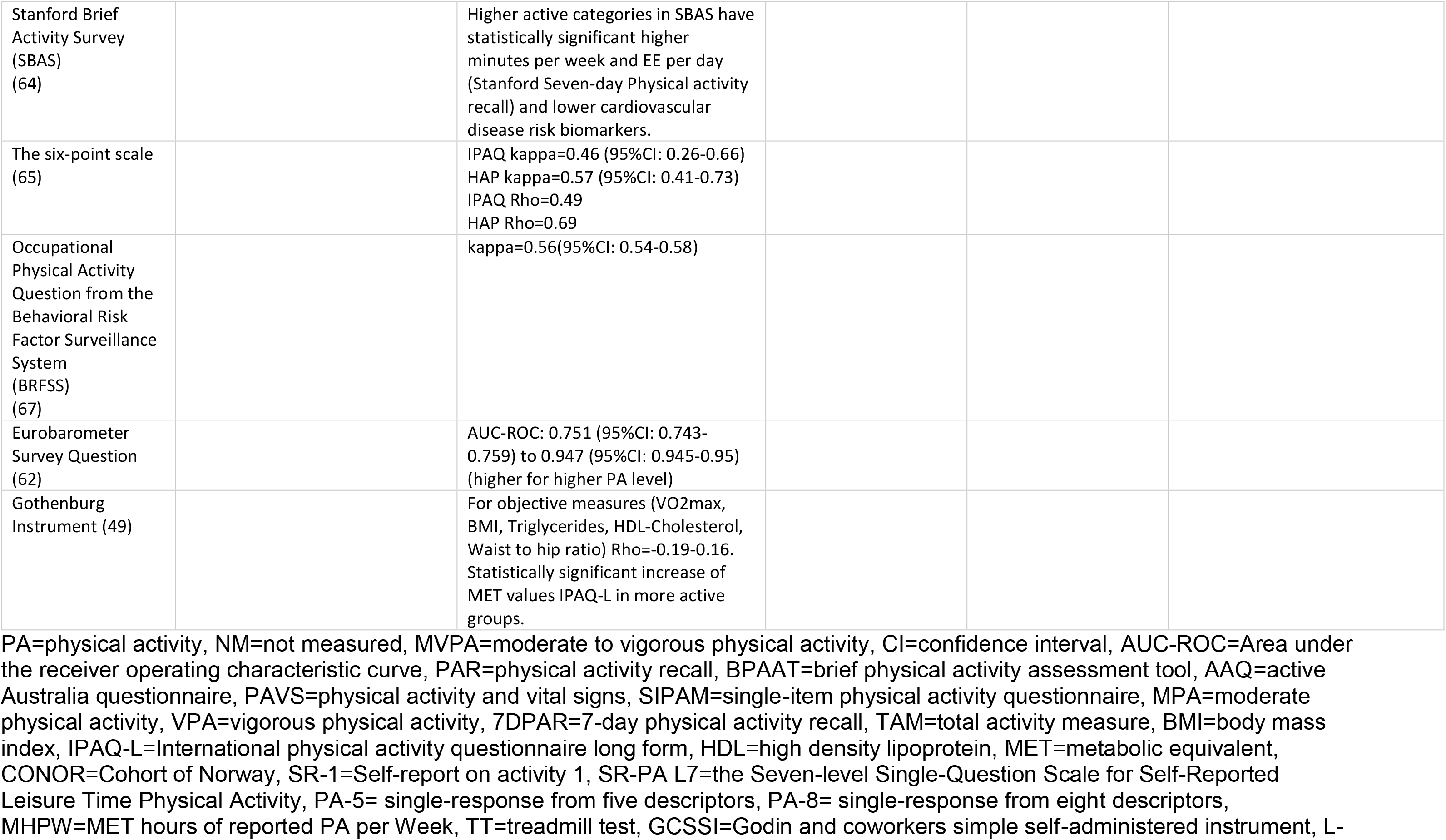
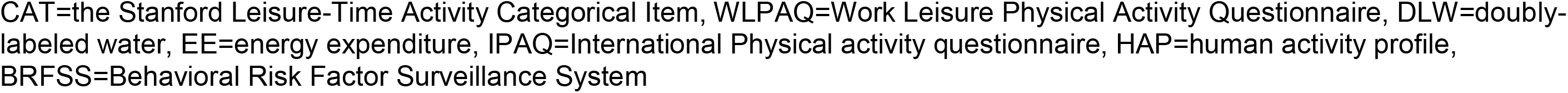
Summary of measurement properties.

Eleven studies measured the reliability of brief PAQs. All of them used a test-retest procedure to measure the consistency of the PAQs. Statistical methods and test-retest intervals varied widely between studies. Overall, studies showed moderate to good reliability levels of the PAQs.

The validity of PAQs was assessed in 33 studies. As a “gold standard” for validation, 13 studies used other PAQs, 15 studies validated brief PAQs against accelerometers or pedometers, and 11 studies compared results of the brief PAQs to other objective measurements, such as BMI (28, 32, 47, 48), VO_2_ max (47, 49), or doubly labeled water (34). Validity coefficients were in general considerably lower than reliability coefficients. The majority of results showed weak validity of brief PAQs against objective measurements and weak to moderate validity against other PAQs.

### Length and complexity of the questionnaires

The texts of all questionnaires and information about their length and readability level are presented in Table 3. It should be noted that, while some PAQs were created and conducted in other languages, length and readability were calculated for the English versions.

**Table 3.**
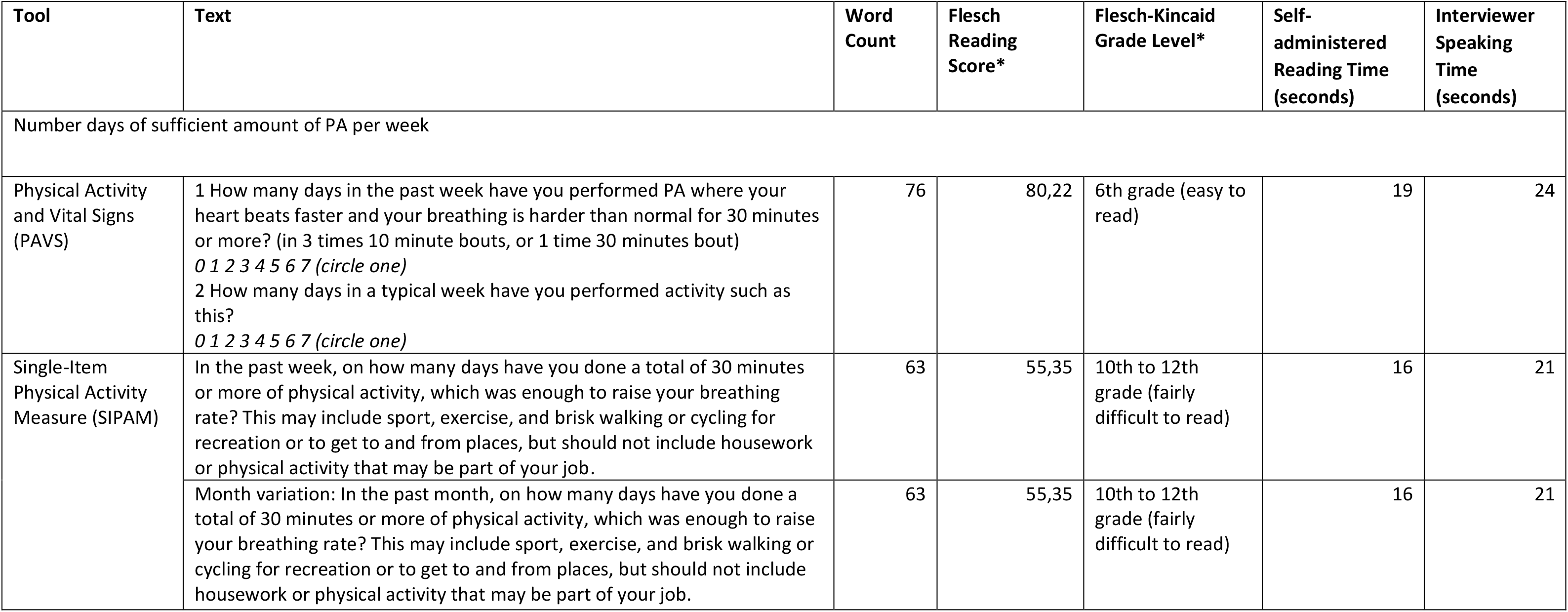
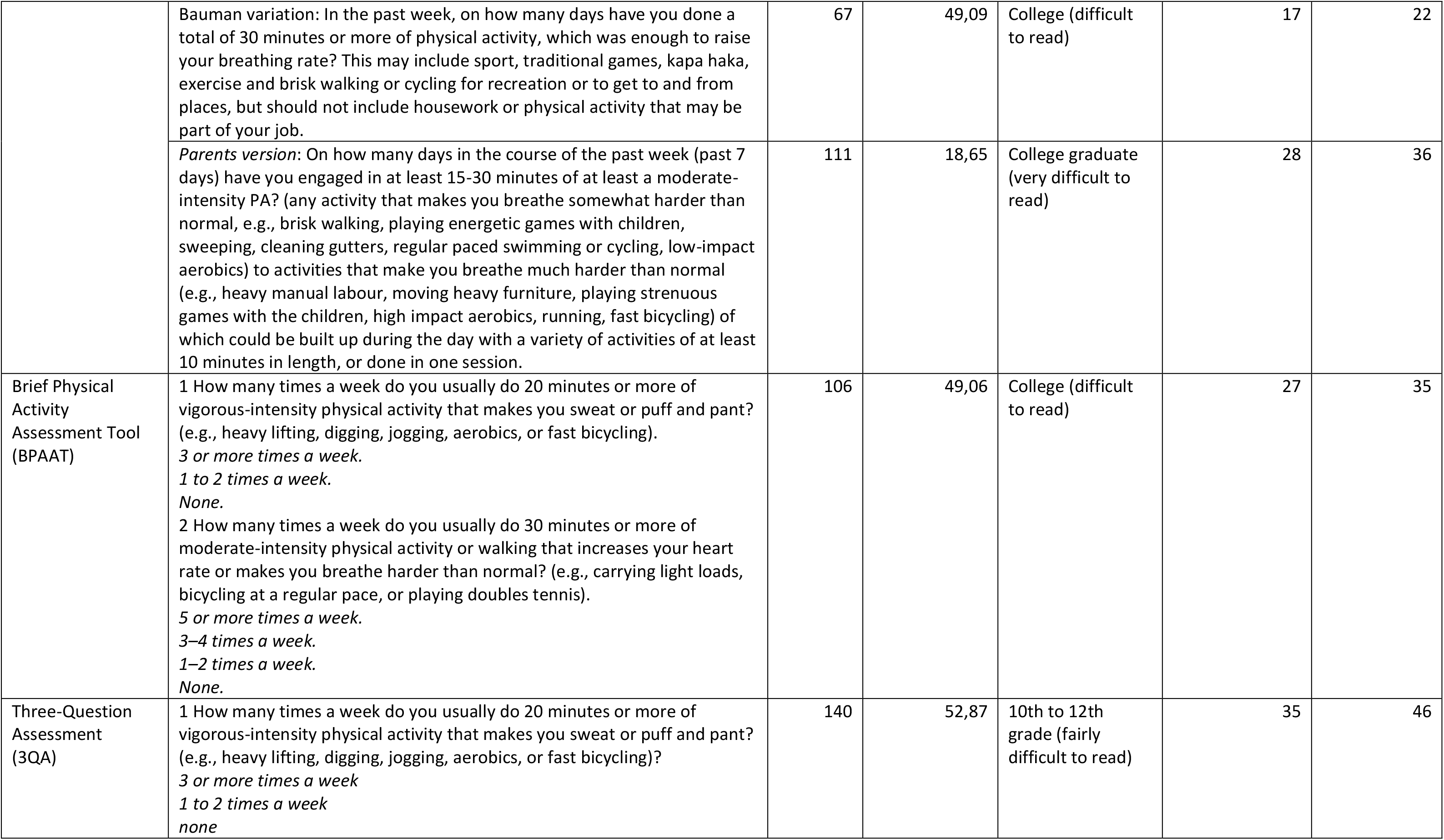
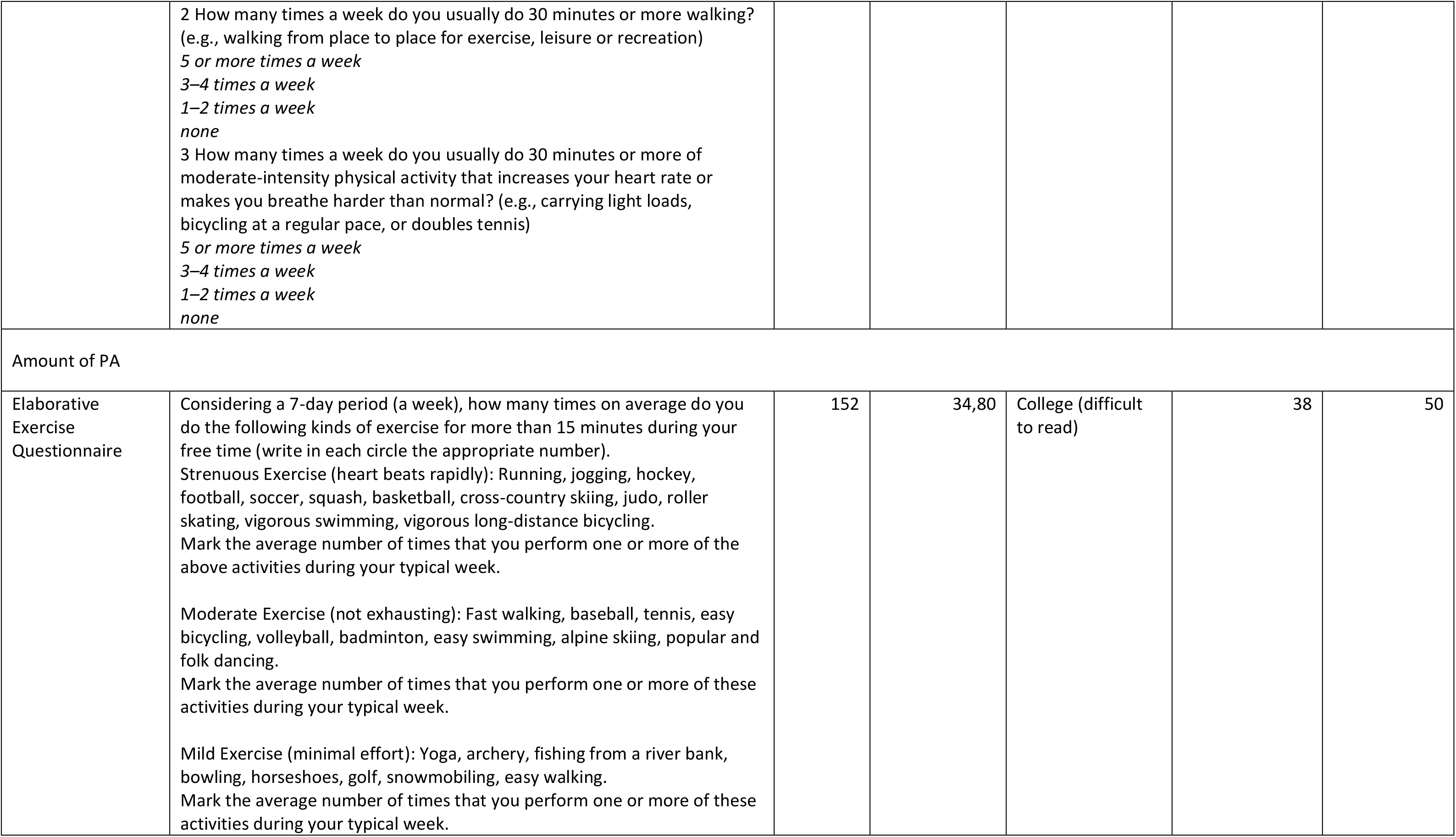
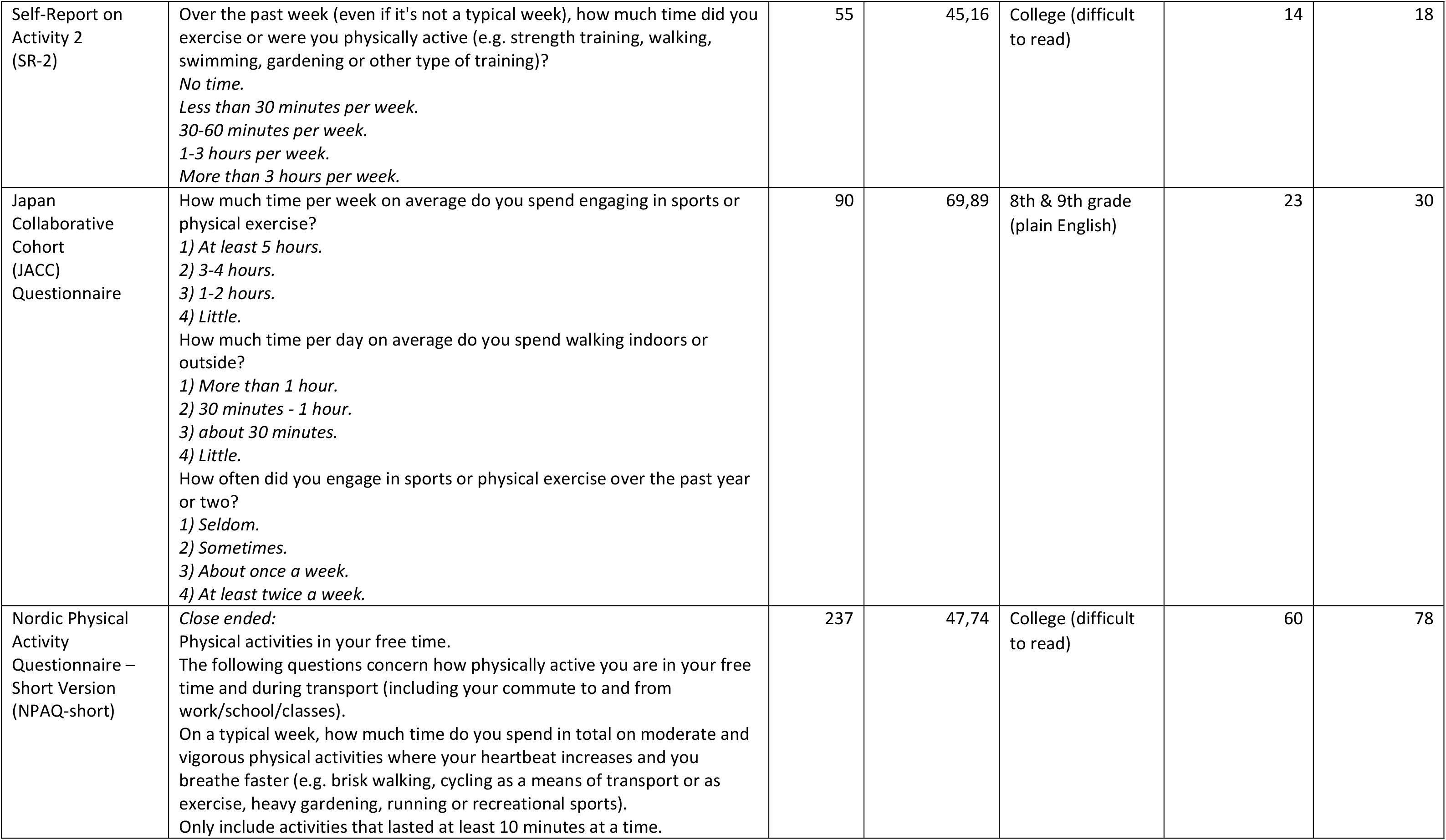
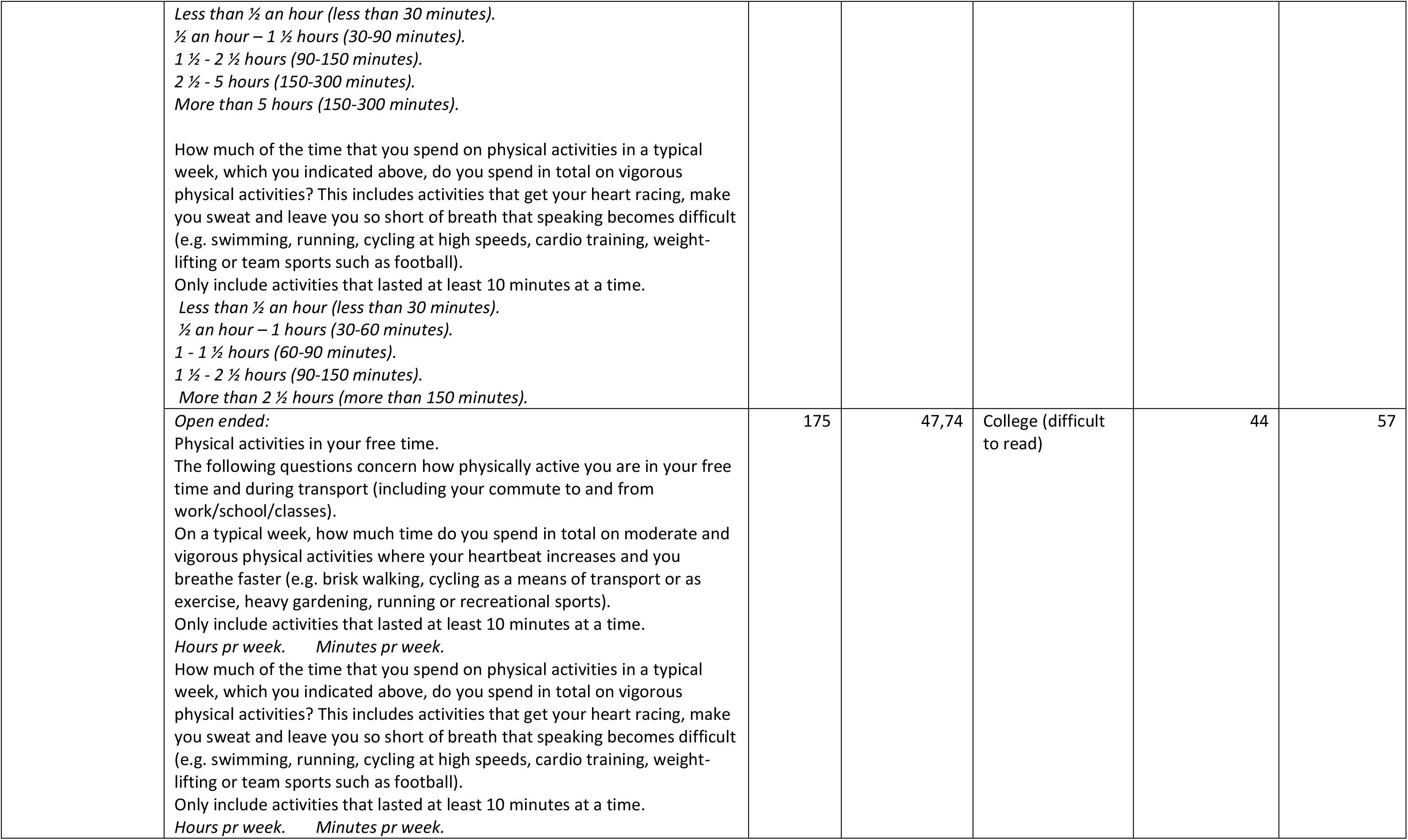
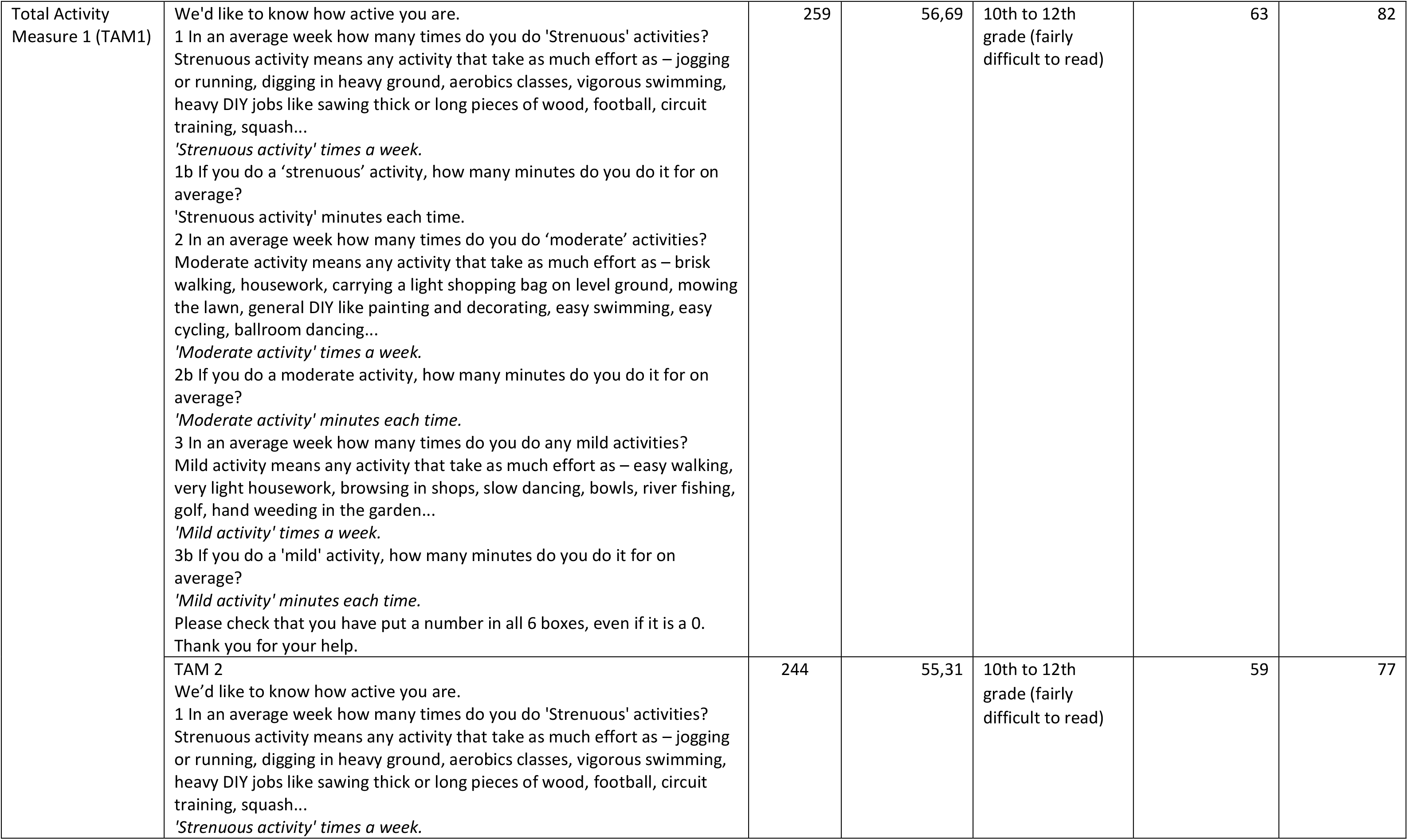
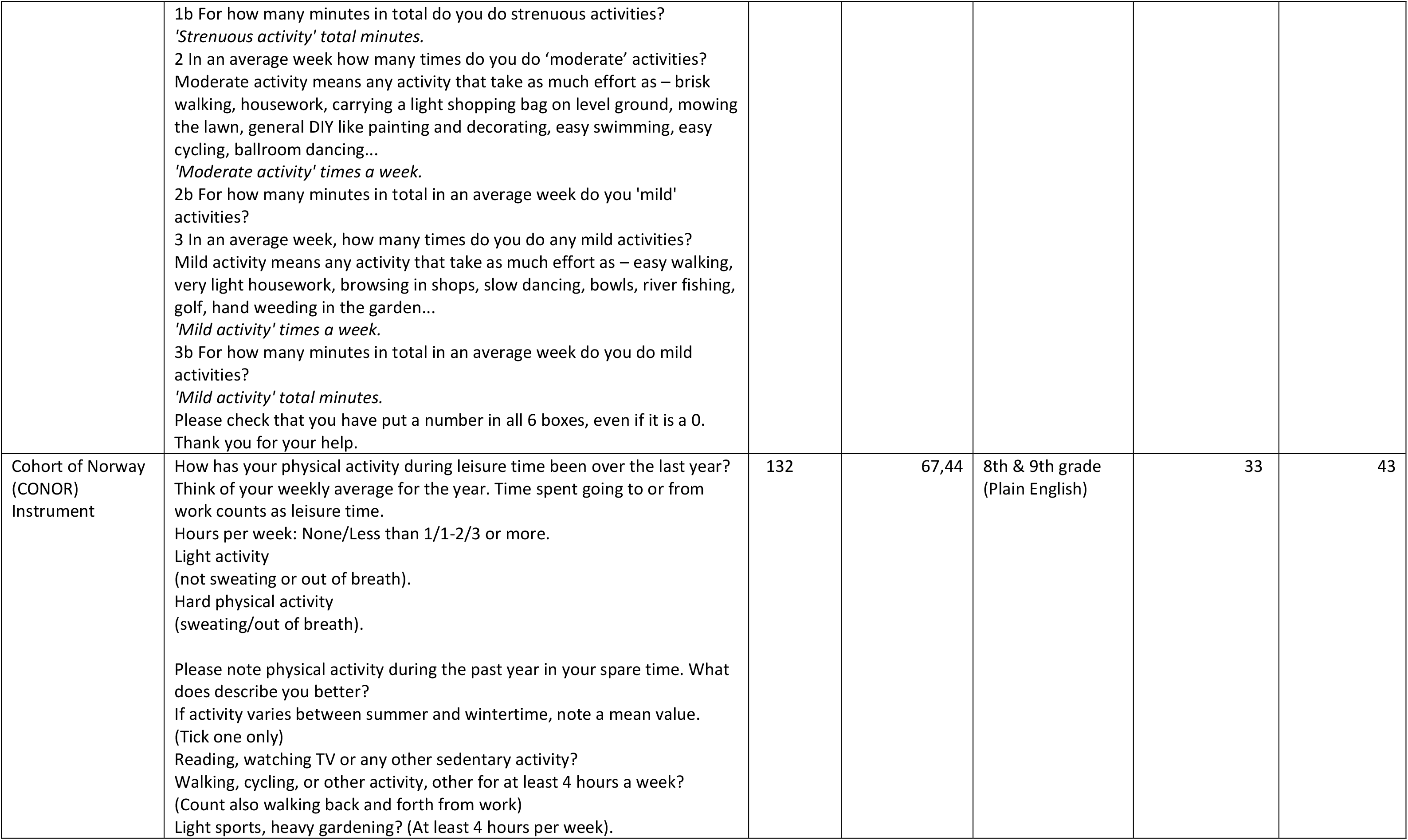
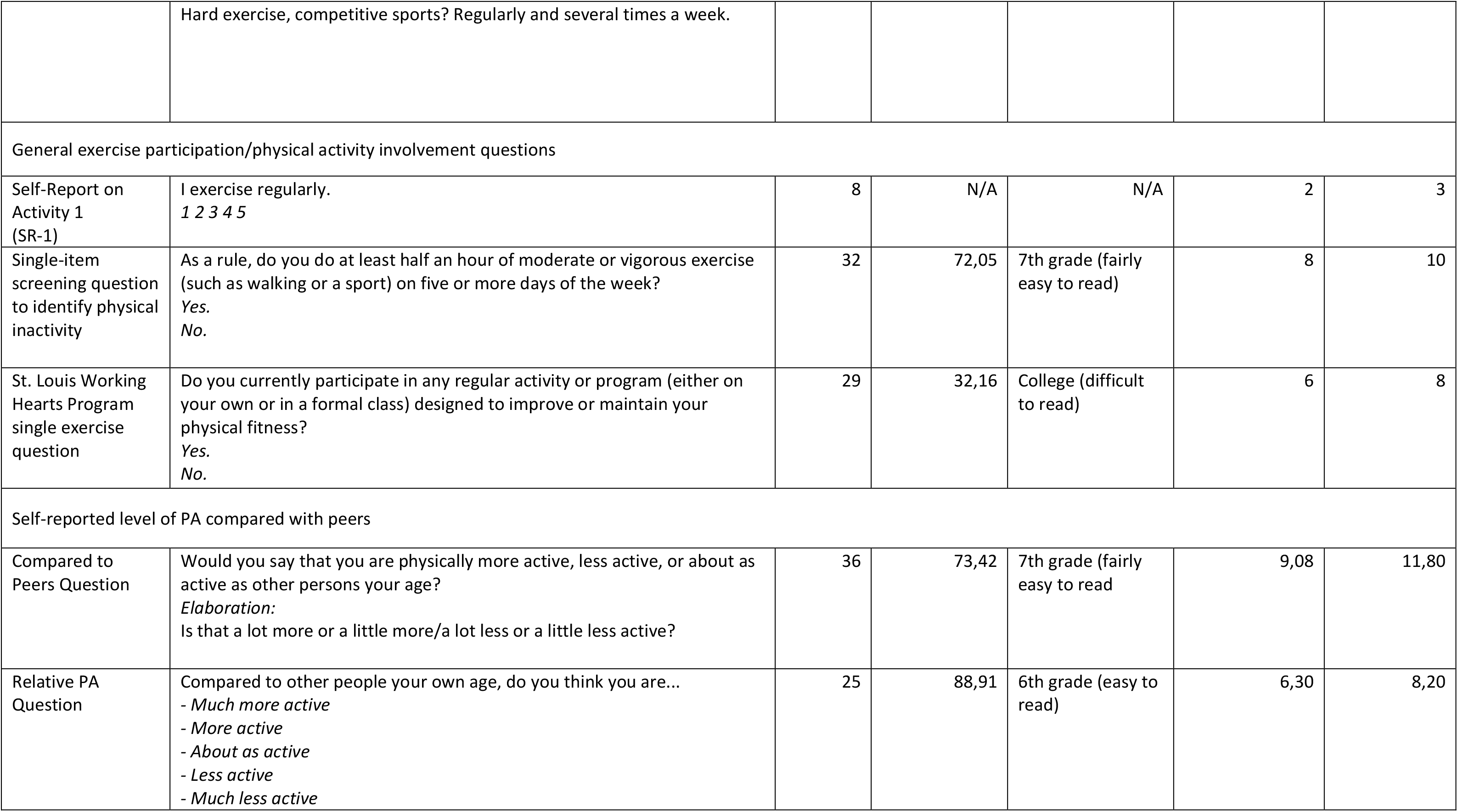
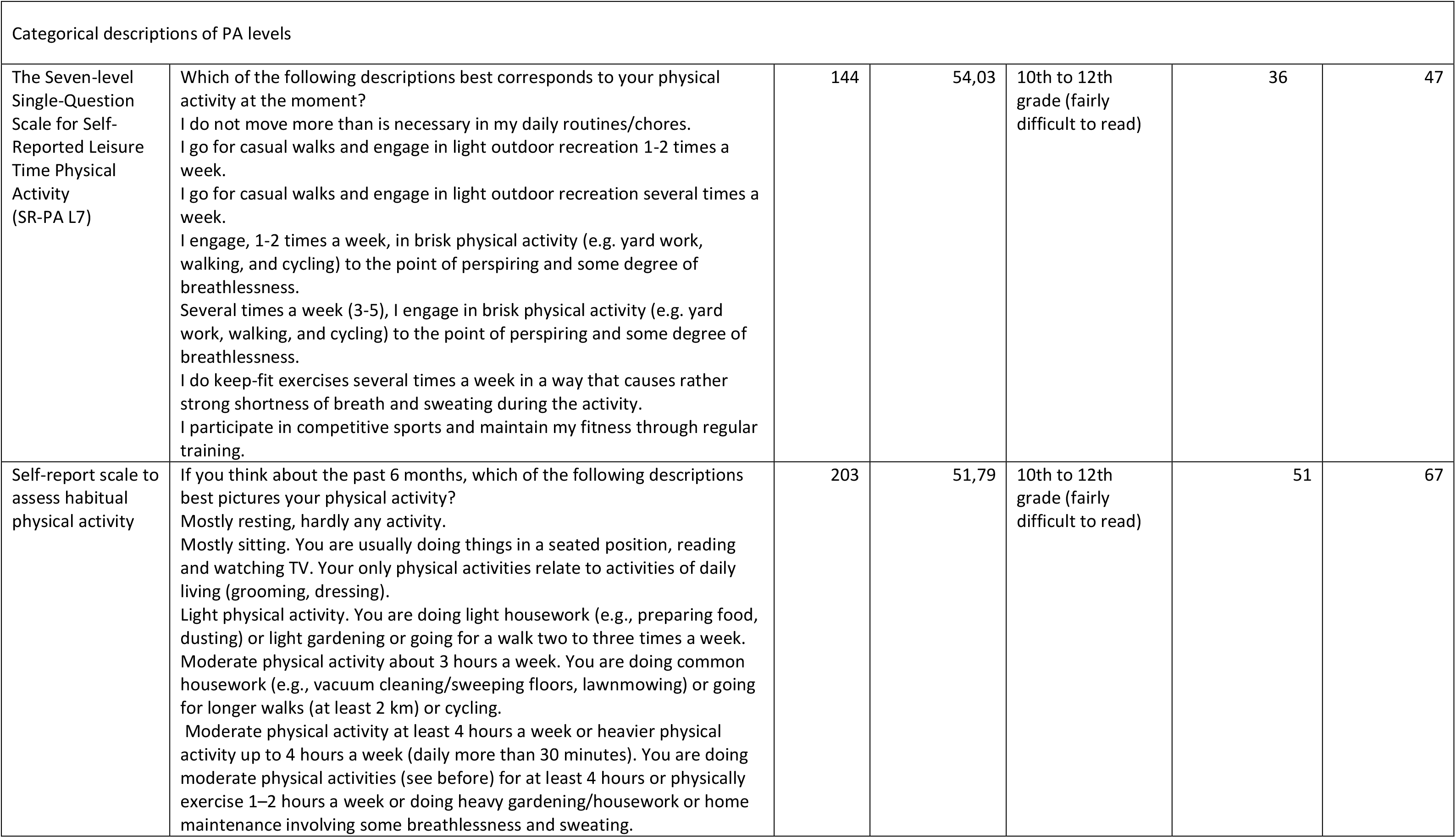
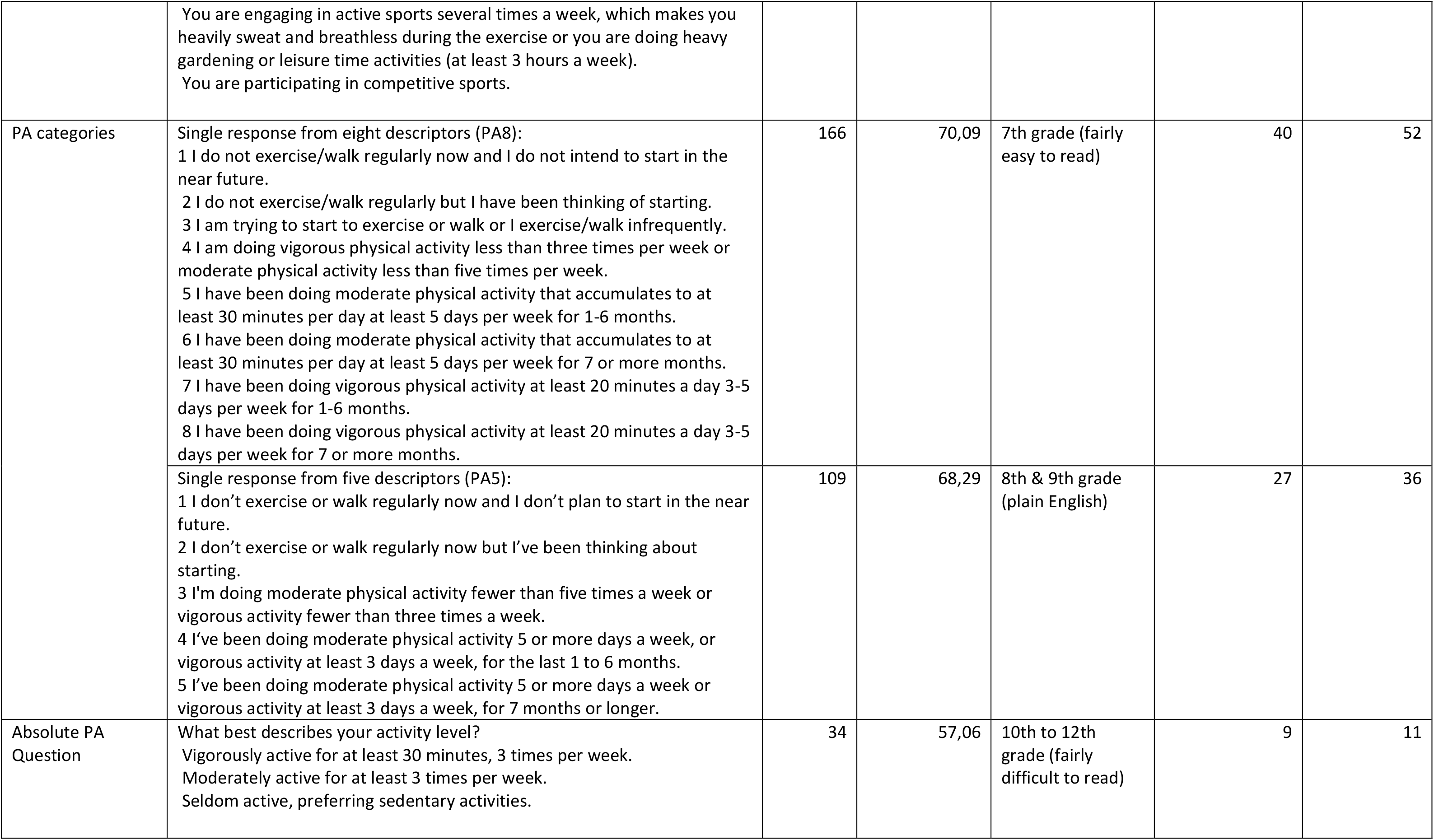
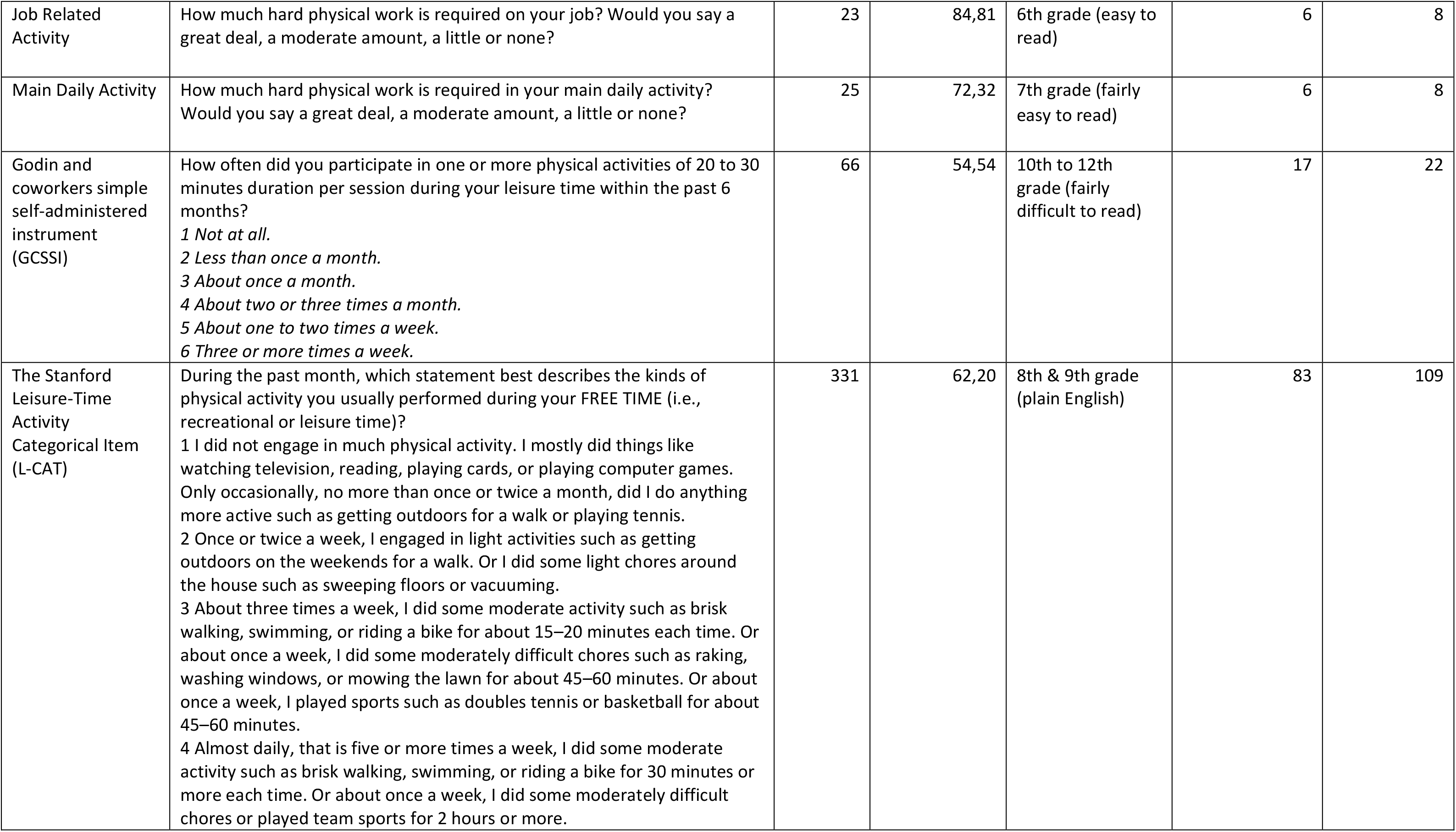
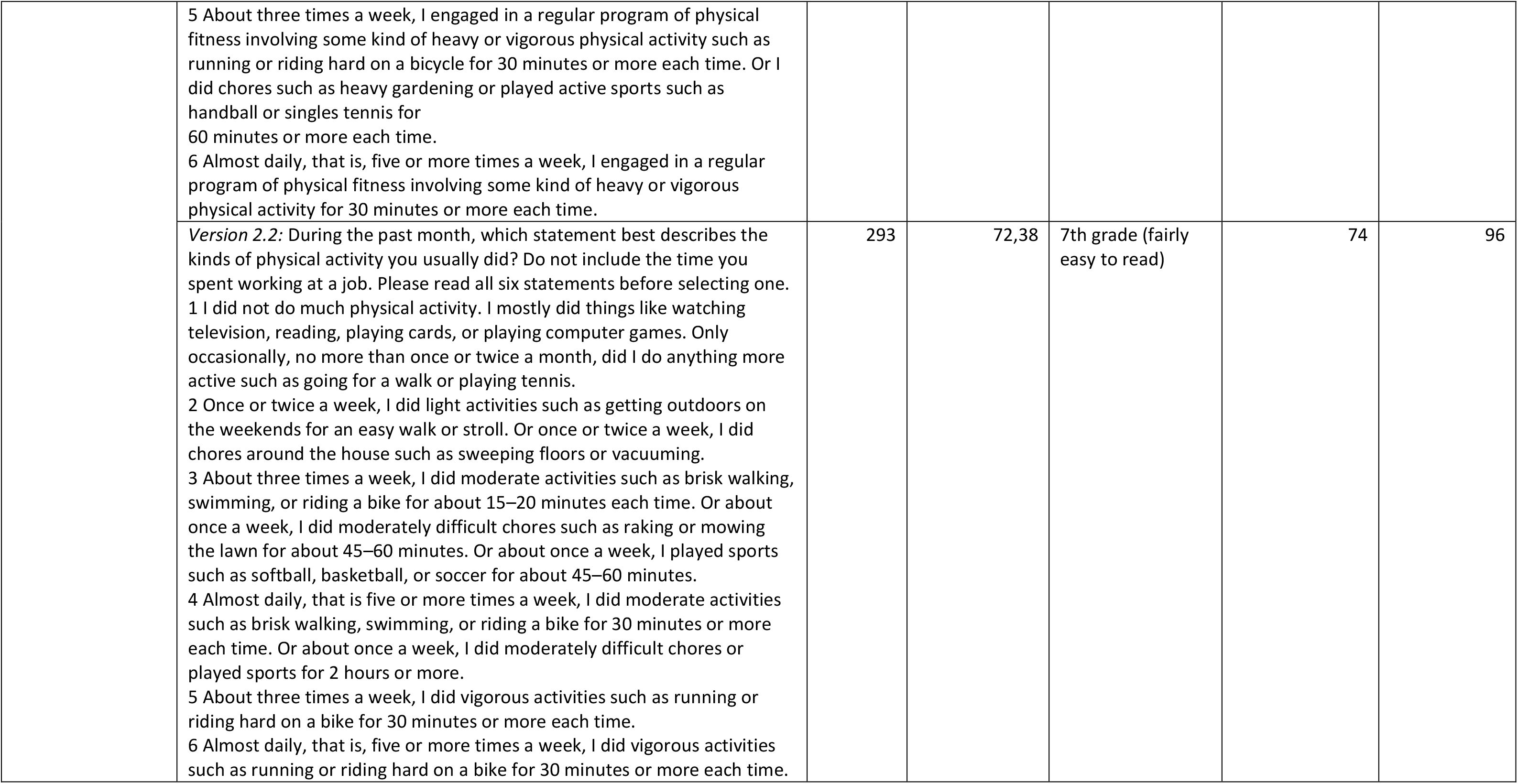
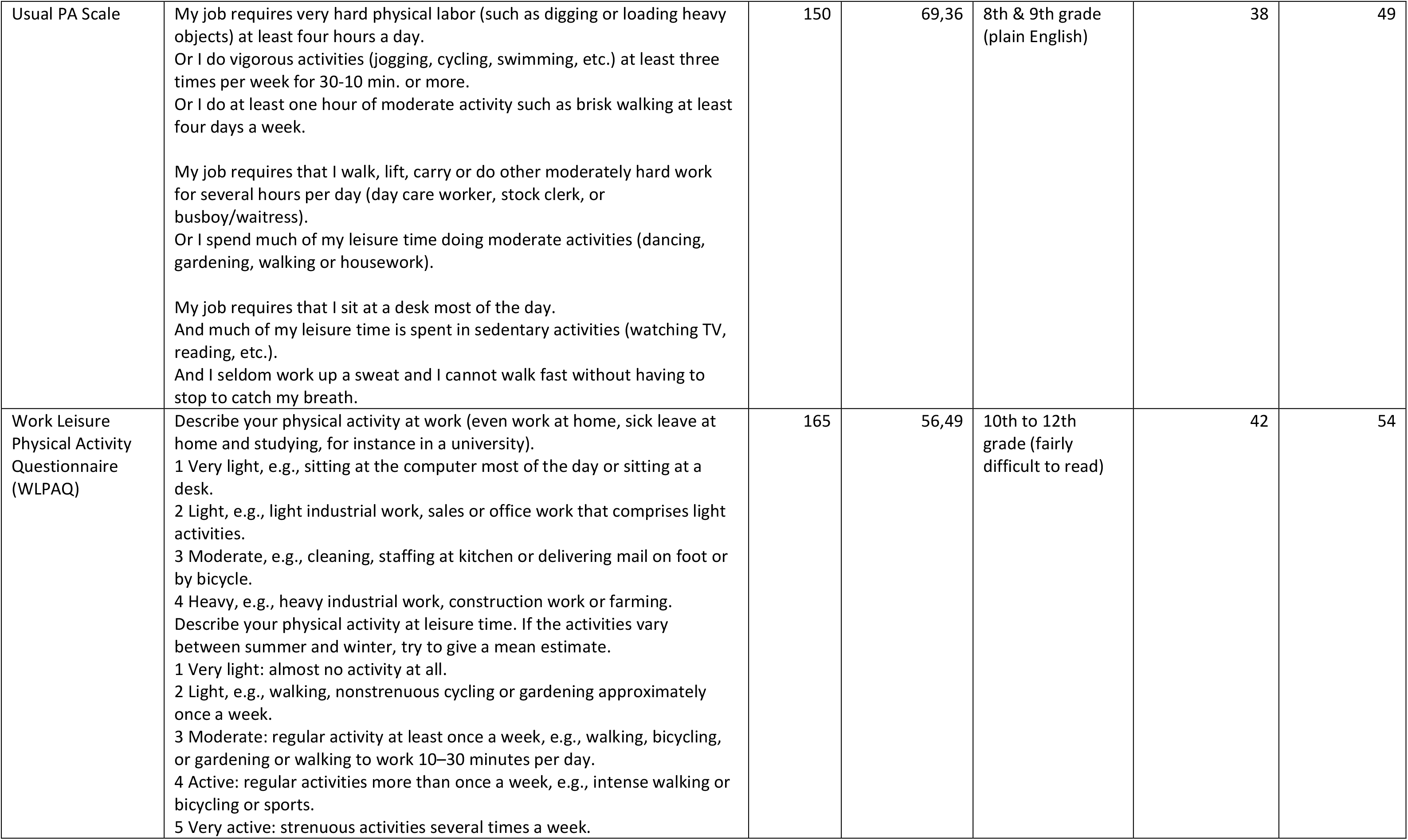
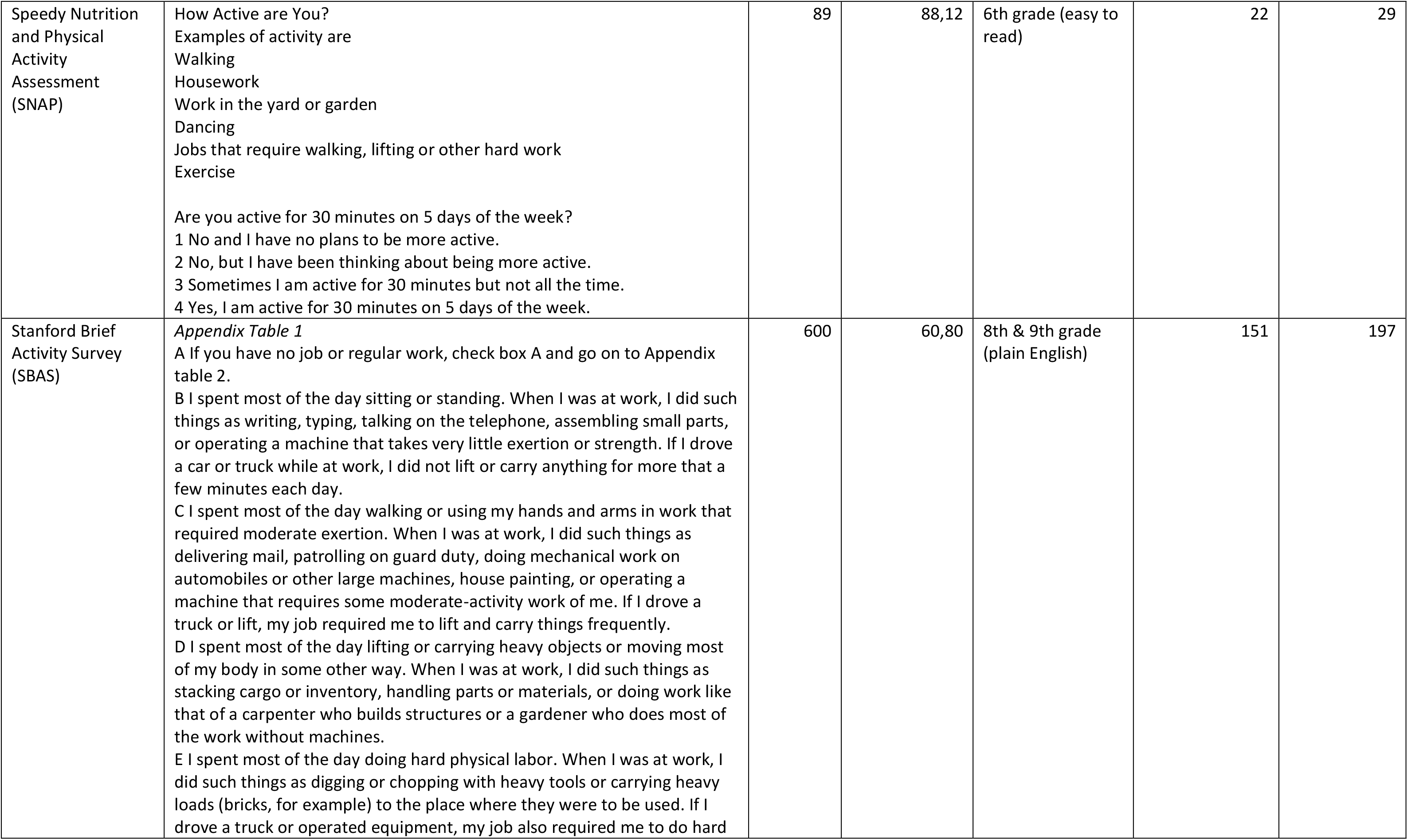
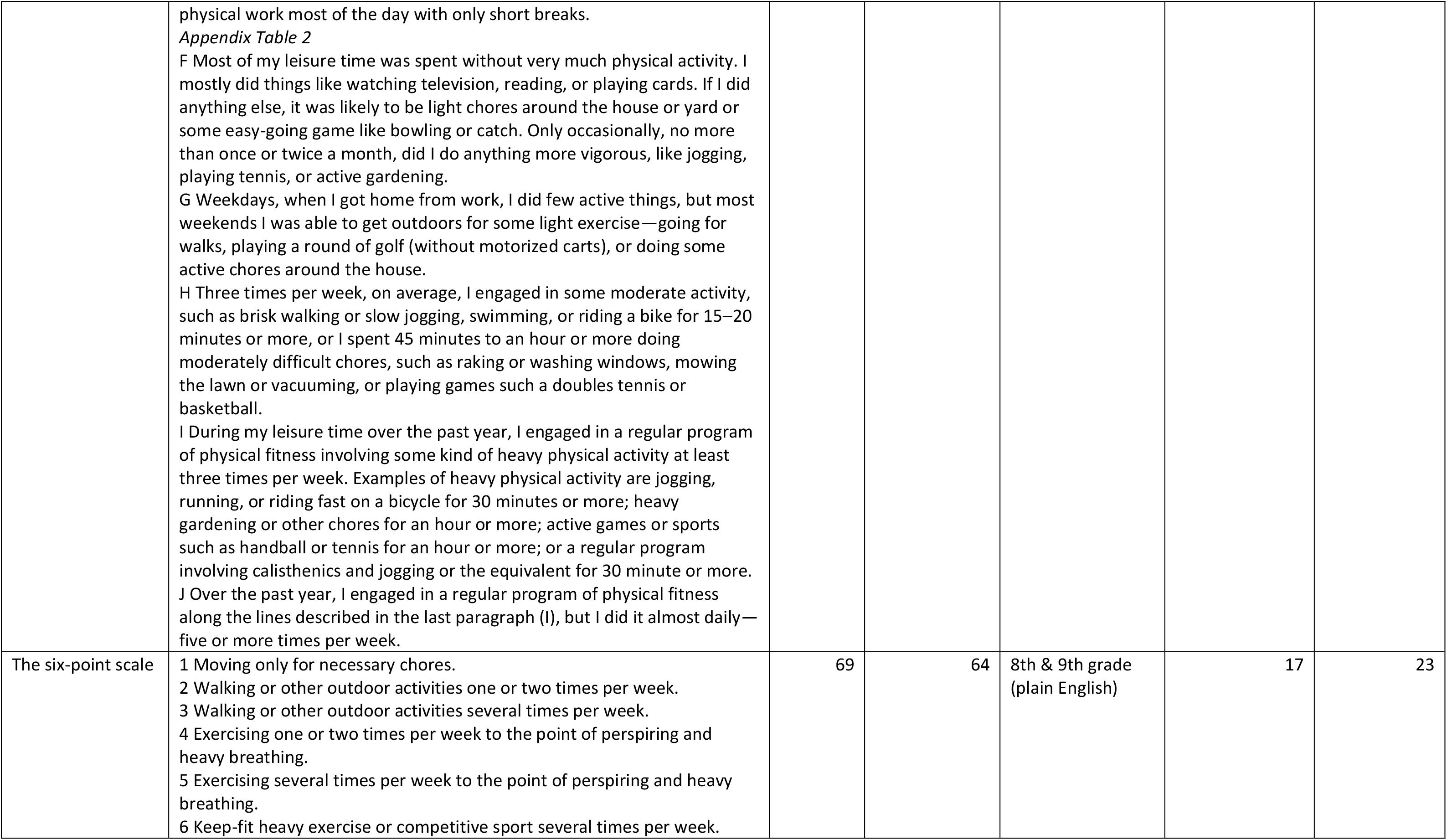
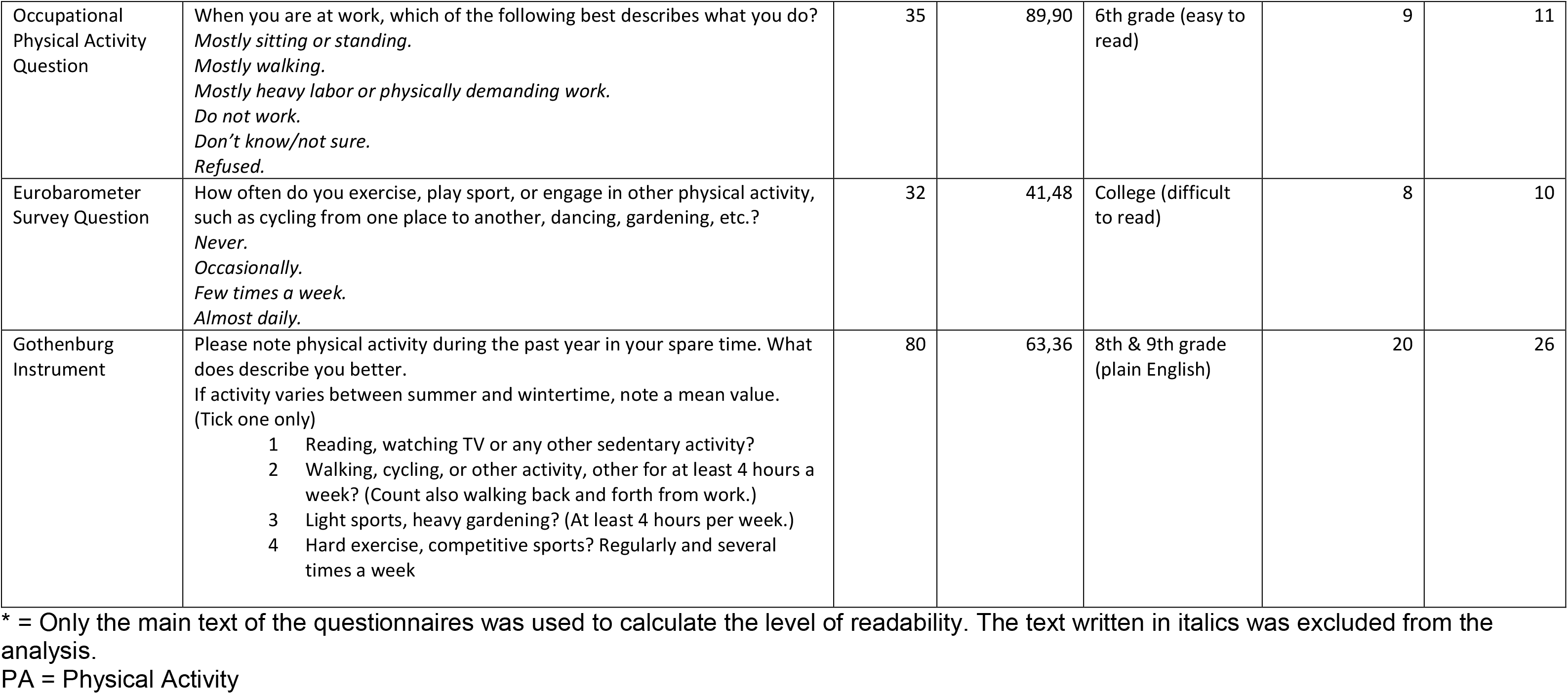
Length and readability levels of short physical activity questionnaires.

Most of the assessed PAQs (n=26) can be read by in silence by the respondent or aloud by the interviewer in less than one minute. However, some questionnaires presented as “brief” by the authors of the identified studies contain a large amount of text and require more than one minute for reading: the Nordic Physical Activity Questionnaire (NPAQ) short, the Total Activity Measure (TAM), the Self-report Scale to Assess Habitual Physical Activity, the Stanford Leisure-Time Activity Categorical Item, and the Stanford Brief Activity Survey.

The calculation of the readability levels with the Flesch-Kincaid formula showed that only 17 out of 31 brief PAQs had readability levels of “easy to read” (n=9) or “plain English,” (n=8) and can be easily understood by the majority of the population (see Figure 2). Other questionnaires have long sentences and/or many complicated words (three syllables and more), making them difficult to read. Seven questionnaires scored as “difficult to read” and require college-level education; it is likely that people with lower education levels will find them hard to understand.

**Figure 2.**
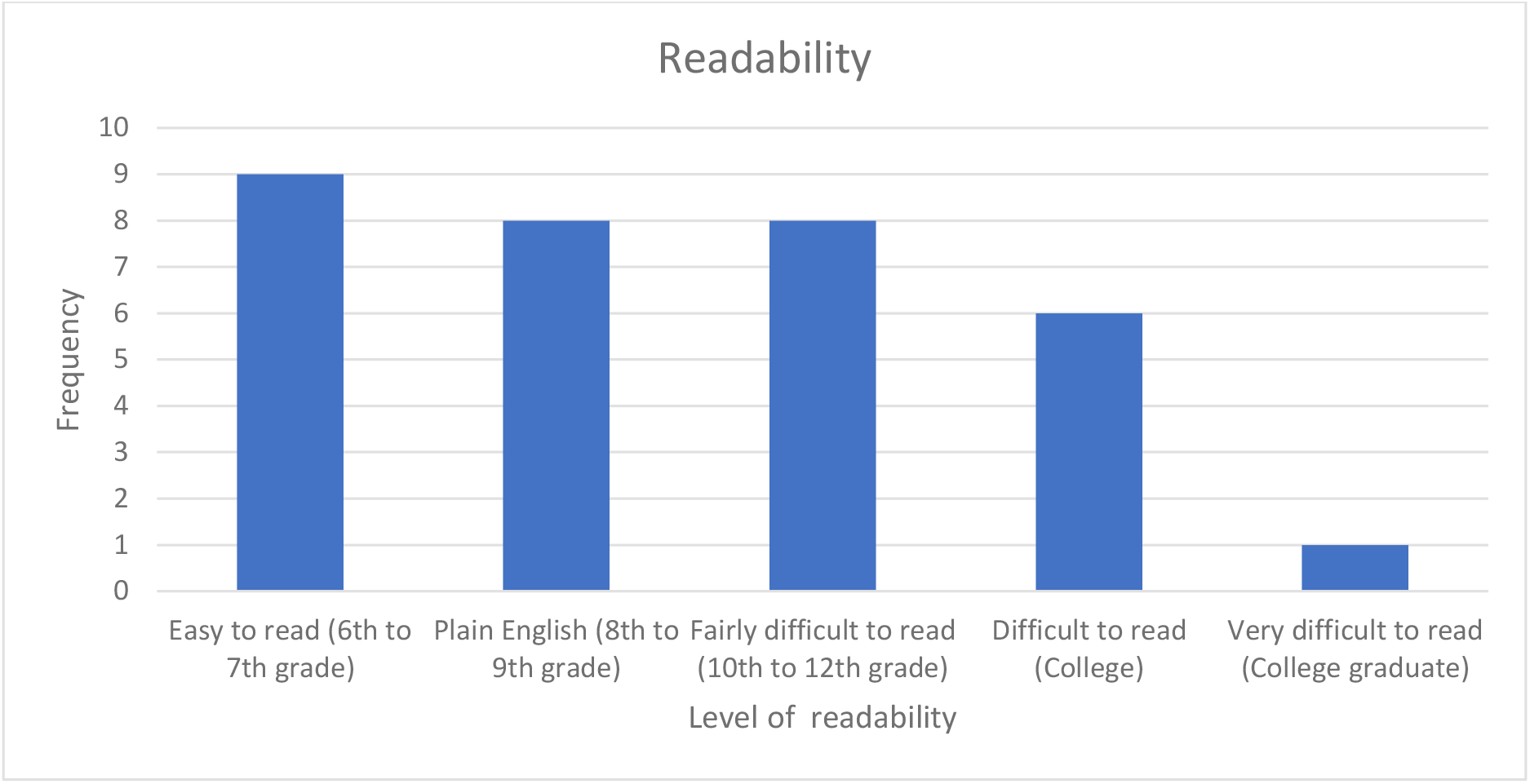
Readability levels for brief PA questionnaires (English language versions).

### Comparison of brief PAQs

Data on both validity and reliability were only available for nine of the 31 assessed PAQs. These brief PAQs were chosen for an in-depth comparison. Their summarized measurement properties and linguistic characteristics are presented in Table 4.

**Table 4.** Comparison of the brief physical activity questionnaires by reliability, validity, length and readability.

| Tool | Reliability | Validity |  | Word count | Readability |
| --- | --- | --- | --- | --- | --- |
|  |  | Against objective measurement | Against other PAQ |  |  |
| Single Item Physical Activity Measure (SIPAM) | Good (0,63-0,82) | Poor to Good (0,22-0,81) | Poor to Good (0,33-0,81) | 63 – original<br>111 – parents version | Fairly difficult to read<br>Parents version - Very difficult to read |
| Brief Physical Activity Assessment Tool (BPAAT) | Moderate to good (0,53-0,72) | Poor to moderate (0,18-0,43) | Moderate to good (0,45-0,64) | 106 | Difficult to read |
| Three-Question Assessment | Moderate to good (0,56-0,63) | Poor (0,24-0,31) | Moderate (0,39-0,43) | 140 | Fairly difficult to read |
| Nordic physical activity questionnaire (NPAQ) - short (open ended) * | Good (0,82) | Poor (0,33) | NM | 237 | Difficult to read |
| Total Activity Measure (Version 2) | Good (0,82) | Poor (0,36-0,38) | NM | 241 | Fairly difficult to read |
| Japan Collaborative Cohort (JACC) Questionnaire* | Moderate (0.39 - 0.56) | NM | Moderate (0.43-0.60) | 90 | Plain English |
| Relative PA Question (Compared to peers) | Moderate (0,56) | Poor to moderate (0,28-0,57) | NM | 25 | Easy to read |
| The Seven-level Single-Question Scale * | Good (0,7) | Poor (0,33) | NM | 144 | Fairly difficult to read |
| Absolute PA Question | Good (0,7) | Poor (0,1-0,33) | NM | 34 | Fairly difficult to read |
| A single-item 5-point rating of usual PA (Usual PA Scale) | Good (0,68-0,88) | NM | Good (0,66) | 147 | Plain English |
| The Stanford Leisure-Time Activity Categorical Item (L-CAT) | Good (0,64-0,8) | Poor (0,36-0,38) | NM | 331 | Plain English |

\* = the study used not the English version of the questionnaire, PAQ=physical activity questionnaire

#### The Single Item Physical Activity Measure (SIPAM)

uses a single question to assess the number of days per week on which 30 minutes or more of PA are performed (excluding housework and work-related PA). For this PAQ, the largest number of studies on validity and reliability were found. The SIPAM showed good reliability levels in all studies, however, results on validity varied considerably between studies – from poor to good – against both objective measurements and other subjective PAQs. Zwolinsky et al. (2015) also measured SIPAM’s ability to identify people that meet or do not meet WHO PA recommendations. The tool showed a low diagnostic capacity compared to the IPAQ. The agreement between the SIPAM and the IPAQ was low for identification of participants who meet WHO PA recommendations (kappa= 0.13, 95% CI 0.12 to 0.14) and moderate for the classification of inactive participants (kappa= 0.45, 95% CI 0.43 to 0.47) (42).

#### The Brief Physical Activity Assessment Tool (BPAAT)

consists of two questions, one regarding the frequency and duration of vigorous PA and the other regarding moderate PA and walking performed in an individual’s usual week. By combining the results of both questions (scores can range from 0 to 8), the subject can be classified as insufficiently (0–3 score) or sufficiently active (≥4 score). Results of reviewed studies showed that the BPAAT has moderate to good levels of reliability and validity in comparison with other PAQs, however comparison with accelerometers identified only poor to moderate validity.

Smith et al. (2005) evaluated the <u>Three-Question Assessment</u> variant of the BPAAT which has separate questions about moderate PA and walking. The results of the study did not find a considerable difference in validity and reliability between the two-and three-question versions. One study also reported that more physicians preferred the two-question version (BPAAT) as it is shorter and therefore easier to use (Smith, 2005).

#### The Nordic Physical Activity Questionnaire-short (NPAQ-short) includes one question

about moderate to vigorous PA and a second question about vigorous PA. Danquah et al. (2018) compared open-ended and closed-ended questions with the open-ended version achieving better results. The open-ended version showed good reliability but performed similarly to other questionnaires, showing poor validity when compared against objective PA measures. The analyses showed that the questionnaire was one of the longest and was rated “difficult to read”. The agreement with accelerometer data in identification of people that meet or do not meet the WHO PA recommendations was low (kappa=0.42).

The <u>Total Activity Measures (TAM)</u> includes three open-ended questions about strenuous, moderate, and mild PA. The revised second version, <u>TAM2,</u> asks about the total time spent at each activity level over a 7-day period. The TAM2 showed good reliability when validated against objectively-measured PA. The word count was high at 241, and readability was rated as “fairly difficult”.

The <u>Japan Collaborative Cohort (JACC) Questionnaire</u> has three items. Two questions focus on leisure-time PA, i.e., time per week engaging in sport or PA (with options ranging from “little” to “at least 5 hours”), and frequency of engagement in sport over the past year (options from “seldom” to “at least twice a week”). The third question asks about daily walking patterns (options from “little” to “more than 1 hour”). The questionnaire showed moderate reliability after one year and moderate validity when compared against a more in-depth interview by a trained researcher. The questions are 90 words in total and were rated as “plain English.”

The <u>Relative PA Question</u> is the shortest of the compared PAQs and has an easy readability level. It allows respondents to compare their level of PA with other people of the same age and to choose from five categories ranging from “much more active” to “much less active.” The questionnaire has a moderate reliability level and showed poor validity against objective measures.

The <u>Absolute PA Question</u> asked participants to choose what best describes their activity level from three options: (1) vigorously active for at least 30 minutes, three times per week; (2) moderately active at least three times per week; or (3) seldom active, preferring sedentary activities. The questionnaire is relatively short, but as most of the other brief PAQs, it showed high reliability and poor validity against objective measures.

The <u>Seven-Level Single-Question Scale (SR-PA L7)</u> requires respondents to assign themselves to a level ranging from “I do not move more than is necessary in my daily routines/chores” and “I participate in competitive sports and maintain my fitness through regular training.” These seven items are supposed to categorize respondents as maintaining low, medium, or high PA. The SR-PA L7 showed poor validity when compared to objectively measured PA but good reliability. The scale is moderately long at 144 words and was rated as “fairly difficult to read.”

#### A single-item 5-point rating of usual PA (Usual PA Scale)

includes descriptions of three PA levels: highly active, moderately active, and inactive. Respondents need to read a description of each category and identify their usual PA level from the list. The questionnaire’s readability level was ranked as “plain English,” and it showed good reliability and validity levels. However, validation was done only against another PAQ which is not well-known and requires further research.

#### The Stanford Leisure-Time Activity Categorical Item (L-CAT)

is a single-item questionnaire that consists of six descriptive PA categories ranging from inactive to very active. Each category consists of one or two statements describing common activity patterns over the past month, differing in frequency, intensity, duration, and types of activity. The categories are described in “plain English,” but this made the questionnaire longer than the other questionnaires. The L-CAT showed high reliability but poor validity against pedometer and accelerometer validation.

## Discussion

This review assessed the validity, reliability, and readability of brief PAQs for adults. To our knowledge, it is the first review focused specifically on brief PAQs and also the first to assess their readability. The sheer number of brief PAQs we identified (n=31) indicates a high research interest in such instruments. However, it also indicates a lack of harmonization when it comes to PA assessment using brief questionnaires.

Overall, most PAQs were reported to have poor or moderate validity. They showed higher validity levels against other self-reported tools than against objective PA measures. Although reliability is an important measurement property, it was assessed only in 11 studies. About half of all PAQs showed a moderate to good level of reliability. These results are in line with other reviews of PAQs (18, 19, 50, 51).

A significant difficulty in conducting this review was that the studies used different methods for validation, varying time-intervals between repeated measurements, and different statistical methods to analyze data. Complete data on validity and reliability were only available for nine PAQs, and only those could be compared in greater detail. All of these factors prevented a more detailed comparison of results across PAQs. In general, the methodological quality of the included studies was modest.

The most common flaws were comparably small sample sizes, a lack of sample size justification, and a poor description of the validity and reliability assessment process. Additionally, most studies utilized convenience samples, making it impossible to assess if measurement properties would differ between adults with different levels of socioeconomic status and/or educational attainment.

Also, the included PAQs used different concepts of measuring PA, further complicating a direct comparison between instruments. For example, the SIPAM measures on how many days per week respondents perform 30 minutes or more of PA; the Absolute PAQ requires respondents to choose from several descriptions of different PA levels; and the Relative PA question asks them to compare their level of PA with peers. Some PAQs, such as the TAM, aim to assess the total volume of moderate-to-vigorous PA. Others focus on particular PA domains. And yet others assess only leisure-time PA. This variety can be partly explained by efforts to keep PAQs short at the price of excluding some PA dimensions (type, duration, intensity, volume). However, it could also be interpreted as a lack of common understanding about which dimensions of PA should be assessed with brief PAQs. It should also be taken into account that none of the reviewed brief PAQs allowed for the measurement of whether a person fulfills the WHO physical activity recommendations (52).

All of the included studies were conducted in highly developed nations, and 23 of them took place in English-speaking countries. This most certainly biased the results, since terminology related to PA differs between languages, as do levels of literacy (53). Ultimately, to assess reliability and validity of PAQs, many more studies should be conducted in developing nations to obtain a more accurate assessment of their suitability for international surveillance systems. Certainly, in order to stimulate such research, funding opportunities need to be made available to research teams from such nations.

The results of this study point out that many PAQs have low readability levels. This is particularly disturbing when considering that most of these PAQs were tested in developed nations with comparatively high levels of literacy. Potentially, poor readability is related to the modest measurement properties that PAQs commonly come with. Questionnaires that are more readable or might even have been co-developed (54) with the intended population groups are likely to yield better measurement properties. This would also strengthen the case of integrating them into PA surveillance systems. However, at this point in time, there is a dearth of research relating PAQs and their measurement properties to readability. Advancing knowledge in this field would also benefit longer PA questionnaires (such as IPAQ and GPAQ) that are widely utilized in surveillance and potentially score low on readability as well.

This review comes with certain limitations. The search was limited to scientific databases, and only studies published in peer reviewed scientific journals were included. This can lead to a publication bias, as all other types of publications and gray literature were excluded. The varying measurement methods and conditions complicated the comparison of findings from different studies and limited data analysis to a narrative description of differences between tools. This led to more subjective results and greater difficulty in identifying the tools with the best measurement properties. It should be also taken into account that the Flesch-Kincaid readability formula was created for longer texts and not adapted for short questionnaires. Consequently, the readability levels of PAQs presented here should be interpreted with caution.

## Conclusion

This systematic review sheds light on the validity, reliability, and readability of short PAQs. Results indicate that additional research is needed, notably regarding their reliability and validity, their readability, and their applicability to non-English speaking and/or developing countries.

Recent years have seen a shift from self-report questionnaires towards objective measures in PA surveillance. This shift has been partially motivated by the persistently modest measurement properties of PAQs, as well as technological advancements in the quality and affordability of accelerometers and other PA measurement tools. However, objective PA assessment comes with its own set of limitations (55), and self-report and objective measures have been described as measuring entirely different parameters. Right now, it seems unclear what the future for PA surveillance might hold and if brief PAQs will continue to play a role.

In this regard, the key merit of short PAQs is that they require substantially less time to complete than the established PA surveillance questionnaires (GPAQ, IPAQ, and EHIS-PAQ). This makes them highly appealing for use in surveillance systems. In order for them to be included in existing systems, more studies would need to investigate how they compare to longer PAQs. Potentially, if their validity and reliability are found to be at a comparable level, we might see surveillance systems gradually shift from longer towards briefer PAQs.

## Declarations

Ethics approval and consent to participate: Not applicable.

### Consent for publication

Not applicable.

Availability of data and materials: Data sharing is not applicable to this article as no datasets were generated or analysed during the current study.

## Authors’ contributions

AT, KA-O and IR conceptualized the study. AT conducted the literature search. AT, RA, RGP, and TA performed abstract/title screening, full-text selection, data extraction, and quality assessment. AT with support from KA-O, SM and PG prepared a draft of the manuscript. All authors critically reviewed the draft and approved the final manuscript.

## Data Availability

All relevant data are within the manuscript and its Supporting Information files.

## Acknowledgements

Not applicable.

## List of abbreviations

BPAAT: Brief Physical Activity Assessment Tool
EHIS-PAQ: European Health Interview Survey Physical Activity Questionnaire
GPAQ: Global Physical Activity Questionnaire
IPAQ: International Physical Activity Questionnaire
JACC: Japan Collaborative Cohort Questionnaire
L-CAT: The Stanford Leisure-Time Activity Categorical Item
NPAQ-short: The Nordic Physical Activity Questionnaire-short
PA: physical activity
PAQ: physical activity questionnaire
SIPAM: Single Item Physical Activity Measure
SR-PA L7: The Seven-level Single-Question Scale for Self-Reported Leisure Time Physical Activity
TAM: Total Activity Measure

